# Wastewater to clinical case (WC) ratio of COVID-19 identifies insufficient clinical testing, onset of new variants of concern and population immunity in urban communities

**DOI:** 10.1101/2022.04.19.22274052

**Authors:** Patrick M. D’Aoust, Xin Tian, Syeda Tasneem Towhid, Amy Xiao, Elisabeth Mercier, Nada Hegazy, Jian-Jun Jia, Shen Wan, Md Pervez Kabir, Wanting Fang, Meghan Fuzzen, Maria Hasing, Minqing Ivy Yang, Jianxian Sun, Julio Plaza-Diaz, Zhihao Zhang, Aaron Cowan, Walaa Eid, Sean Stephenson, Mark R. Servos, Matthew J. Wade, Alex E. MacKenzie, Hui Peng, Elizabeth A. Edwards, Xiao-Li Pang, Eric J. Alm, Tyson E. Graber, Robert Delatolla

**Affiliations:** Department of Civil Engineering, University of Ottawa, Ottawa, Canada; Children’s Hospital of Eastern Ontario Research Institute, Ottawa, Canada; Department of Biology, University of Waterloo, Waterloo, Canada; Department of Chemical Engineering and Applied Chemistry, University of Toronto, Toronto, Canada; Department of Chemistry, University of Toronto, Toronto, Canada; Department of Laboratory Medicine and Pathology, University of Alberta, Edmonton, Canada; Department of Biological Engineering, Massachusetts Institute of Technology, Cambridge, Massachusetts, United States of America; Data, Analytics and Surveillance Group, UK Health Security Agency, London, United Kingdom

**Keywords:** COVID-19, SARS-CoV-2, variant, wastewater-based epidemiology, wastewater surveillance, public-health

## Abstract

Clinical testing has been the cornerstone of public health monitoring and infection control efforts in communities throughout the COVID-19 pandemic. With the extant and anticipated reduction of clinical testing as the disease moves into an endemic state, SARS-CoV-2 wastewater surveillance (WWS) is likely to have greater value as an important diagnostic tool to inform public health. As the widespread adoption of WWS is relatively new at the scale employed for COVID-19, interpretation of data, including the relationship to clinical cases, has yet to be standardized. An in-depth analysis of the metrics derived from WWS is required for public health units/agencies to interpret and utilize WWS-acquired data effectively and efficiently. In this study, the SARS-CoV-2 wastewater signal to clinical cases (WC) ratio was investigated across seven different cities in Canada over periods ranging from 8 to 21 months. Significant increases in the WC ratio occurred when clinical testing eligibility was modified to appointment-only testing, identifying a period of insufficient clinical testing in these communities. The WC ratio decreased significantly during the emergence of the Alpha variant of concern (VOC) in a relatively non-immunized community’s wastewater (40-60% allelic proportion), while a more muted decrease in the WC ratio signaled the emergence of the Delta VOC in a relatively well-immunized community’s wastewater (40-60% allelic proportion). Finally, a rapid and significant decrease in the WC ratio signaled the emergence of the Omicron VOC, likely because of the variant’s greater effectiveness at evading immunity, leading to a significant number of new reported clinical cases, even when vaccine-induced community immunity was high. The WC ratio, used as an additional monitoring metric, complements clinical case counts and wastewater signals as individual metrics in its ability to identify important epidemiological occurrences, adding value to WWS as a diagnostic technology during the COVID-19 pandemic and likely for future pandemics.

## 1 Introduction

Severe acute respiratory syndrome coronavirus 2 (SARS-CoV-2) has had profound effects around the world, claiming millions of lives and significantly impacting public health since its initial identification in December 2019 (World Health Organization, 2021). The virus is an enveloped, positive-sense RNA virus and, like other coronaviruses, predominantly causes respiratory and intestinal tract infections (Chen et al., 2020; Xu et al., 2020; Zou et al., 2020). SARS-CoV-2 infections in humans can cause acute respiratory illness leading to elevated mortality and morbidity rates (Challen et al., 2021; Fajnzylber et al., 2020; O’Driscoll et al., 2021). Early in the COVID-19 pandemic, molecular diagnostic assays based on the reverse transcription quantitative polymerase chain reaction (RT-qPCR) technology were rapidly developed (Zhu et al., 2020) and became the principal testing method used to determine if patients were infected with COVID-19. Other testing methodologies and technologies, such as rapid antigen testing and LAMP RT-PCR, were also used globally. However, RT-qPCR testing remains the gold standard due to its sensitivity and specificity, particularly at lower viral loads (Hirotsu et al., 2020; Uhteg et al., 2020). Clinical testing in most jurisdictions around the world, including Canada, has predominantly targeted symptomatic individuals (Long et al., 2020; Oran and Topol, 2020; Rivett et al., 2020). The incubation period of SARS-CoV-2, together with the inherent lag associated with the RT-qPCR clinical testing and reporting pipeline (typically measured in days) generates a significant delay between infection incidence and the reporting of results, potentially delaying life-saving public health actions. Wastewater surveillance (WWS) has emerged as an exemplary early indicator of the incidence and disease burden of SARS-CoV-2 in the community (Al Huraimel et al., 2020; Cao and Francis, 2021; Cavany et al., 2022; D’Aoust et al., 2021a). In the first year of the pandemic, research on the pathogenicity of SARS-CoV-2 revealed that fecal shedding of viral RNA is frequent, where approximately 30-90% of cases feature fecal shedding (Fontana et al., 2020; Hua et al., 2020; Huang et al., 2020; Jones et al., 2020; Ling et al., 2020; Santos et al., 2020). Thus, community-level measurements of the viral signal in wastewater are thought to act as a proxy for community SARS-CoV-2 burden (Bibby et al., 2021a; D’Aoust et al., 2021a; Karthikeyan et al., 2021; La Rosa et al., 2020; Olesen et al., 2021; Quilliam et al., 2020). In addition, following wastewater-based viral signal over time provides near real-time information on the virus dynamics such as incidence, prevalence, and disease burden (Ahmed et al., 2020; Bivins et al., 2020; Chakraborty et al., 2021; D’Aoust et al., 2021b; Medema et al., 2020; Peccia et al., 2020; Randazzo et al., 2020; Sherchan et al., 2020; Wu et al., 2020). Furthermore, sequencing of wastewater or variant-specific RT-qPCR assays can also detect and track the emergence/re-emergence of variants of interest/variants under investigation/variants of concern (VOI/VUI/VOC) (Graber et al., 2021; Lee et al., 2021; Peterson et al., 2021; Yaniv et al., 2021).

With the growing amount of data available from multiple locations and COVID-19 resurgences, there now exists an opportunity to explore the relationships between clinical reporting and WWS results to better understand and interpret patterns of both. With this data, there is clear potential for hypothesis-driven approaches to identify mechanisms underlying relationships between clinical testing patterns and observed SARS-CoV-2 viral signal in wastewater within and between communities and facilities (Bibby et al., 2021b). It is hypothesized that analyzing the change in the intrinsic relationship existing between clinical testing and observed SARS-CoV-2 viral signal may reveal additional information regarding testing strategy effectiveness in approximating true disease incidence in a community. Xiao *et al*. proposed a metric to relate wastewater to epidemiological information; the wastewater to clinical (WC) ratio (Xiao *et al.,* 2022). It was proposed by Xiao *et al*. (2022) that tracking and analyzing changes in the WC ratio across time could yield additional information regarding clinical testing adequacy in approximating true disease incidence, and perhaps ease of spread of a pathogen or its severity in the community.

In this study, the WC ratio was analyzed over periods of 8 to 21 months in the seven Canadian cities/regions of Ottawa, Kitchener, Toronto, Peel, Waterloo, Calgary, and Edmonton to help identify if it could identify occurrences of major epidemiological importance during the COVID-19 pandemic. The major epidemiological events were as follows: i) a period of insufficient clinical testing during a change in clinical testing strategy (i.e., walk-in testing changed to appointment-based testing); ii) the emergence of a more infectious VOC (Alpha) in the context of low vaccine- or naturally-acquired community immunity; iii) the emergence of a more transmissible and virulent VOC (Delta) in the context of high community immunity, and iv) the emergence of a more transmissible VOC (Omicron) in the context of immunity escape in a susceptible community. Furthermore, other possible confounding factors such as the age, demographics, and vaccination rate of reported new clinical cases were also considered to assure observed trends were not affected by these elements. The objective of this study was therefore to investigate and quantify the ability of the WC ratio to identify major epidemiological occurrences and further elucidate the relationship existing between WWS and reported clinical cases to complement and enhance the value of WWS and its enhance its usefulness as an applied diagnostic technology.

## 2 Materials and methods

Analytical testing completed in this study occurred in four different laboratories throughout Canada, and as such specific methodologies, site-specific sampling schedules, sample processing, extraction, and quantification techniques differ slightly between the various cities/regions. Before starting, all employed methodologies were compared as part of a broader pan-Canadian interlaboratory study (Chik et al., 2021) and determined to be of similar performance under the tested conditions. A list of the testing periods included in this study and the major epidemiological events identified at each testing site is shown below in Table 1.

**Table 1:** Testing periods and analyzed major epidemiological events identified per studied site.

| Location | Studied period | Identified change in clinical testing strategy changed to appointment based only | Identified onset of allelic proportionality of B.1.1.7 (Alpha) VOC reaching 40-60% | Identified onset of allelic proportionality of B.1.617.2 (Delta) VOC reaching 40-60% |
| --- | --- | --- | --- | --- |
| Ottawa | 06-04-2020 to 09-01-2022 | 06-10-2020 | 23-03-2021 | 31-07-2021 |
| Toronto | 11-02-2021 to 20-11-2021 | - | 02-03-2021 | 22-06-2021 |
| Kitchener | 01-12-2020 to 15-09-2021 | - | 10-03-2021 | 06-06-2021 |
| Peel | 20-09-2020 to 30-08-2021 | 06-10-2020 | 22-02-2021 | 28-05-2021 |
| Waterloo | 07-12-2020 to 15-09-2021 | - | 27-03-2021 | 14-06-2021 |
| Edmonton | 12-07-2020 to 26-09-2021 | - | 23-02-2021 | 18-07-2021 |
| Calgary | 09-07-2020 to 21-09-2021 | - | 22-03-2021 | 18-07-2021 |

Furthermore, a summary of the characteristics of each water resource recovery facility (WRRF) is shown in Supplementary Material (Table S1). The studied data sets for most locations stopped in August 2021, however, the studied data set in Ottawa (most frequently sampled and tested) was chosen to be extended into the Omicron VOC onset.

### 2.1 Sample collection, enrichment, concentration, and extraction

#### Ottawa

24-hour composite samples of primary clarified sludge were collected from the City of Ottawa’s sole WRRF, the Robert O. Pickard Environmental Centre (ROPEC), starting on Apr. 8^th^, 2020, with daily sample collection beginning on Sep. 10^th^, 2020. The WRRF services approximately 91% of households and individuals in the City of Ottawa (pop.: 1.1M). Composites were collected every morning and kept on ice until transported to the University of Ottawa for analysis. In this study, samples were analyzed from Apr. 8^th^, 2020, until Jan. 5^th^, 2022. Primary sludge samples were first homogenized well by mixing and then 40 mL of each sludge sample was concentrated by centrifugation for 45 minutes at 10,000 × g. The supernatant was discarded and the wet pellet was centrifuged a second time at 10,000 × g for an additional 5 minutes. The resulting pellet was then homogenized, and 0.250 g ± 0.05 g of the resulting solids pellet was extracted as previously described using Qiagen’s RNeasy PowerMicrobiome Kit (Qiagen, Germantown, MD) using a QIACube Connect automated extraction platform (D’Aoust et al., 2021a, 2021b). RNA was finally eluted in 100 µL of RNase-free water.

#### Toronto

24-hour composite samples of post-grit influent were collected from three water resource recovery facilities in the City of Toronto, including Ashbridge’s Bay (TAB), Highland Creek (THC), and North Toronto (TNT) at frequencies of three to five times per week between Feb. 11^th^, 2021, and Nov. 22^nd^, 2021. After arriving at the laboratory at the University of Toronto, two pseudo-biological replicates (80 mL for TAB and THC samples, and 120 mL for TNT samples) were centrifuged. The wastewater samples were then transferred to 50 mL centrifuge tubes in 40 mL aliquots and centrifuged at 10,000 × g for 45 minutes at 4°C. After removal of the supernatant, the remaining wet pellet was transferred to a 2 mL screw-cap microcentrifuge tube and further centrifuged at 13,000 × g for 1 minute. After completely decanting the supernatant, the final pellet and microcentrifuge tube was weighed. For pellets not exceeding 250 mg, RNA extraction was performed as previously described using Qiagen’s RNeasy PowerMicrobiome Kit (Qiagen, Germantown, MD) (D’Aoust et al., 2021a, 2021b)., with the following modifications to the protocol: 10 µL beta-mercaptoethanol and 100 µL phenol:chloroform:isoamyl alcohol (25:24:1 v/v) mixtures (Invitrogen, CAT# 15593031, USA) were added to the bead beating tube along with the lysis buffer in the kit. For pellets exceeding 250 mg, each pellet was split into two portions to proceed with cell lysis and inhibition removal (IRS), and only half of the combined supernatant from the IRS step was used in the remaining extraction steps. Whole process controls were included for each extraction. RNA was finally eluted in 100 µL of RNase-free water.

#### Kitchener, Peel, and Waterloo

24-hour composite samples of post-grit influent were collected from the following locations: (Kitchener) Region of Waterloo’s water resource recovery facility (Dec. 7^th^, 2020, to Sep. 15^th^, 2021), (Peel) Clarkson water resource recovery facility (Aug. 8^th^, 2020, to Sep. 15^th^, 2021) and G.E. Booth water resource recovery facility (Jul. 20^th^, 2020, to Aug. 30^th^, 2021), and (Waterloo) Waterloo water resource recovery facility (Dec. 7^th^, 2020, to Sep. 15^th^, 2021). Samples were collected at a frequency of three to five times per week. Samples were aliquoted into 250 mL HPDE bottles (Systems Plus, Baden, Canada) and transported on ice to the University of Waterloo. Upon arrival, the sample bottles were well mixed, and a 40 mL aliquot was poured into 50 mL centrifuge tubes containing 4 g PEG-8000 and 0.9 g NaCl and mixed on an orbital shaker at 4°C for 2 h before being stored at 4°C overnight. The samples were then centrifuged at 12,000 × g at 4°C for 90 minutes with no deceleration/brake applied. The majority of the supernatant was discarded, and the remaining sample was centrifuged again at 12,000 × g at 4°C for 5 minutes with no deceleration/brake applied. The remaining supernatant was decanted/pipetted out and the wet weight of the resulting pellet was recorded. Up to 250 mg of the pellet was used for RNA extraction using the RNeasy PowerMicrobiome Kit (Qiagen, Germantown, MD) with the addition of 100 µL of TRIzol (Thermo Fisher, Ottawa, Canada) to the pellet before bead beating. RNA was finally eluted in 100 µL of RNase-free water.

#### Edmonton and Calgary

24-hour composite post-grit wastewater influent samples were collected from the following locations: (Edmonton) EPCOR Gold Bar Wastewater Treatment (May 18^th^, 2020, to Sep. 26^th^, 2021) and the city of Calgary water resource recovery facility (May 17^th^, 2020, to Sep. 21^st^, 2021). Samples were collected at a frequency of three to five times per week. Wastewater concentration and RNA extraction were performed as previously described (Qiu et al., 2021). Briefly, 100 ml wastewater samples were spiked with 100 µl of a suspension of human coronavirus (HCoV) strain 229E (10^5^ IU/ml titrated by TCID50) to monitor virus recovery. The spiked samples were adjusted to a pH of 9.6-10 using 5N NaOH, mixed vigorously for 30 seconds, and then centrifuged at 4,500 × g for 10 min to pellet solids. The liquid fraction was then transferred into a new container, adjusted to pH 7-7.5 using 1.2N HCl, and then concentrated by ultrafiltration using a Centricon Plus-70™ filter with a pore size or Nominal Molecular Weight Limit (NMWL) of 30 KDa (Merck Millipore, Carrigtwohill, Ireland) at 3000 × g for 10 minutes at 4°C. The concentrated sample was adjusted to a final volume of 1 ml by adding phosphate-buffered saline (PBS) and stored at −70°C until RNA extraction. Viral RNA was extracted from ultrafiltration wastewater concentrates using the MagMAX™-96 Viral RNA isolation kit (Thermo Fisher Scientific, Ottawa, Canada) and the King Fisher™ Flex Purification System (Thermo Fisher Scientific, Ottawa, Canada) using 400 µl of wastewater concentrate as input. RNA was finally eluted in 100 µL of elution buffer.

### 2.2 RT-qPCR quantification of SARS-CoV-2, Pepper Mild Mottle Virus, Alpha, Delta, and Omicron VOCs in wastewater

Singleplex RT-qPCR quantification of SARS-CoV-2 targets and Pepper Mild Mottle Virus (PMMoV) was performed in the four laboratories analyzing samples for this study. A summary of the targeted SARS-CoV-2 genes, RT-qPCR reaction characteristics, and QA/QC controls utilized in each laboratory during this study are shown below in Table 2. All probes and primers utilized in this study are included in the Supplemental Material (Table S2). For viral signal normalization, the copies of SARS-CoV-2 (N1/N2) detected per reaction were normalized against the quantified copies of PMMoV from corresponding samples.

**Table 2:** Summary of the RT-qPCR quantification of SARS-CoV-2 targets and PMMoV in all four laboratories which performed analyses for this study.

|  | Ottawa | Toronto | Kitchener, Peel, and Waterloo | Edmonton and Calgary |
| --- | --- | --- | --- | --- |
| **Targeted SARS-CoV-2 regions** | N1, N2, ND3L (Alpha), D63G (Delta), P13L (Omicron) | N1, N2, ND3L (Alpha), D63G/N200 (Delta) | N1, N2, ND3L (Alpha), D63G/N200 (Delta) | N1, N2, E, ND3L (Alpha), D63G (Delta) |
| **Targeted biological normalizer** | PMMoV | PMMoV | PMMoV | PMMoV |
| **RNA template volume (µL)** | 3 (SARS-CoV-2 targets) / 1.5 (PMMoV) | 4 | 5 | 5 |
| **Total RT-qPCR reaction volume (µL)** | 10 | 10 | 20 | 10 |
| **Supermix used** | TaqMan™ 1-Step Fast Virus Master Mix (ABI) | TaqMan™ 1-Step Fast Virus Master Mix (ABI) | TaqPath™ 1-Step RT-qPCR Master Mix (ABI) | TaqMan™ 1-Step Fast Virus Master Mix (ABI) |
| **Standard utilized** | Exact Diagnostics SARS-CoV-2 RNA standard (COV019) | Custom concatenated plasmid containing N1, N2, and PMMoV gene regions | Exact Diagnostics SARS-CoV-2 RNA standard (COV019) | Custom in-vitro transcribed gblock |
| **Standard curve points** | 5-6 | 5-6 | 5-6 | 5-6 |
| **Endogenous spike in** | VSV | - | HCoV-229E, zebrafish dsDNA, MS2 | - |
| **Corrected for recovery efficiency?** | No | No | No | No |
| **Inhibition check?** | Yes, via dilution | Yes, via dilution | Yes, via zebrafish dsDNA | Yes, via dilution |
| **Assay limit of detection (ALOD) for N1, N2, PMMoV targets (copies/reaction)** | 3.2 (N1), 8.1 (N2), 7.6 (PMMoV) | 1.6 (N1), 1.6 (N2) | - | 1.6 (N1), 1.6 (N2) |
| **RT-qPCR conditions** | (All targets)<br>Reverse trans.: 5 min. @ 50°C, 1 cycle<br>Initial denat.: 20 sec. @ 95°C, 1 cycle<br>Denaturation: 3 sec. @ 95°C, 45 cycles<br>Anneal/ext.: 30 sec. @ 60°C, 45 cycles | (All targets)<br>Reverse trans.: 5 min. @ 50°C, 1 cycle<br>Initial denat.: 20 sec. @ 95°C, 1 cycle<br>Denaturation: 3 sec. @ 95°C, 45 cycles<br>Anneal/ext.: 30 sec. @ 60°C, 45 cycles | (N2, HCoV-229E, MS2)<br>Reverse trans.: 15 min. @ 50°C, 1 cycle<br>Initial denat.: 2 min. @ 95°C, 1 cycle<br>Denaturation: 3 sec. @ 95°C, 45 cycles<br>Anneal/ext.: 60 sec. @ 60°C, 45 cycles<br>(N1, PMMoV)<br>Reverse trans.: 15 min. @ 50°C, 1 cycle<br>Initial denat.: 2 min. @ 95°C, 1 cycle<br>Denaturation: 3 sec. @ 95°C, 45 cycles<br>Anneal/ext.: 60 sec. @ 60°C, 45 cycles | (N1, N2, E and ND3L)<br>Reverse trans.: 5 min. @ 50°C, 1 cycle<br>Initial denat.: 20 sec. @ 95°C, 1 cycle<br>Denaturation: 3 sec. @ 95°C, 45 cycles<br>Anneal/ext.: 30 sec. @ 60°C, 45 cycles<br>(D63G)<br>Reverse trans.: 5 min. @ 50°C, 1 cycle<br>Initial denat.: 20 sec. @ 95°C, 1 cycle<br>Denaturation: 3 sec. @ 95°C, 45 cycles<br>Anneal/ext.: 30 sec. @ 55°C, 45 cycles |
| **Primer and probe concentrations** | 500 µM (primers)<br>125 µM (probes) | 500 µM (primers)<br>125 µM (probes) | 500 µM (primers)<br>125 µM (probes) | 800 nM (primers) and 200 nM (probes) (N1, N2, E)<br>900 nM (primers) and 250 nM (probe) (ND3L)<br>800 nM (primers) and 250 nM (probes) (D63G) |
| **Replicate exclusion threshold** | Ct ≥ 0.5 | Ct ≥ 0.5 | Ct ≥ 0.5 | None |
| **Standard curve acceptance characteristics** | $R^2 \geq 0.95$,<br>90% ≤ Eff. ≤ 120% | $R^2 \geq 0.99$,<br>95% ≤ Eff. ≤ 105% | $R^2 \geq 0.95$,<br>95% ≤ Eff. ≤ 105% | External standard curve characteristics (data from 10 separate RT-qPCR runs):<br>$R^2 \geq 0.99$<br>95% ≤ Eff. ≤ 103% (N1 and N2)<br>$R^2 \geq 0.99$<br>92% ≤ Eff. ≤ 113% (ND3L)<br>$R^2 \geq 0.99$<br>90% ≤ Eff. ≤ 109% (D63G) |
| **Other QA/QC performed** | Extraction blank, no extrapolation of values, negative controls | Extraction blank, negative controls, positive controls | Extraction blank, no extrapolation of values, negative controls, positive controls, no RT controls | Extraction blank, negative controls. External standard curve was verified for each RT-qPCR run at two different standard dilutions; Ct values differences between standards in each run vs those from external standard curve should be within ± 1 |

### 2.3 Epidemiological data

Daily epidemiological data including new daily testing, reported positive COVID-19 cases, and demographics of new infections were obtained throughout the study via Ottawa Public Health’s online portal (Ottawa Public Health, 2021), Public Health Ontario’s COVID-19 online data tool (Public Health Ontario, 2021), and the COVID-19 surveillance database of the Government of Alberta (Government of Alberta, 2021). The epidemiological data were provided per geographical area by the various public health units (known in certain jurisdictions as public health agencies) to best match the sewershed of each studied location. stratified by geographic areas or sites having wastewater-based SARS-CoV-2 surveillance included in this study. Vaccination information was obtained via Public Health Ontario’s COVID-19 online data tool (Public Health Ontario, 2021) for locations in Ontario, and from the Government of Canada’s COVID-19 vaccination coverage information page (Government of Canada, 2021).

### 2.4 Statistical analysis

The Pearson’s product-moment correlation coefficient (Pearson’s r) was calculated to evaluate the strength of the relationship between WWS-acquired, PMMoV normalized SARS-CoV-2 viral signal and reported clinical cases throughout the study. Furthermore, to establish if changes occurred in the WC ratio values during key epidemiological occurrences, the Mann-Whitney U test was performed on the 15-day periods preceding and following the key epidemiological occurrences. A non-parametric test (Mann-Whitney U test) was employed to establish significance in changes of the WC ratio during key epidemiological occurrences to remove distribution effects. A regression analysis and subsequent analysis of the residuals were also performed between the 7-day midpoint average daily new reported clinical cases and the 7-day midpoint average observed viral signal in wastewater to establish if any outliers existed.

### 2.5 Calculation of the WC ratio

The WC ratio in this study was calculated by using the PMMoV-normalized SARS-CoV-2 viral signal (N1-N2 average copies/copies PMMoV) and dividing this value by the number of COVID-19 clinical cases on the same day, similar to what was described previously in the literature (Xiao et al., 2022). No lead-lag correction was applied to the calculations of the WC ratio (Cavany et al., 2022; D’Aoust et al., 2021a; Galani et al., 2022; Olesen et al., 2021).

### 2.6 Calculation of VOC allelic proportions

The allelic proportions of different SARS-CoV-2 VOCs were calculated by dividing measured VOC-associated viral copies/g by the universal/non-mutated equivalent genomic region viral copies/g, to obtain a percentage/proportion, as previously described (Graber et al., 2021). This calculation allows to track what percentage of the viral signal observed in wastewater is attributable to the presence of circulating VOCs.

### 2.7 Standardization of SARS-CoV-2 measurements for comparison of curves

The viral signal was first standardized to be of similar scale (on the same axis) by multiplying the PMMoV-normalized viral signal measurements in Ottawa and Toronto by 100,000, Kitchener by 300,000, Waterloo by 500,000, not modifying the measurement of Peel, and dividing the measurements of Edmonton and Calgary by 1,000. This was performed to facilitate initial comparison of results by community.

## 3 Results and discussion

### 3.1 Overview of SARS-CoV-2 surveillance at studied sites

Surveillance of SARS-CoV-2 viral signal at the studied sites began on different dates due to the different laboratories employed to support WWS across the study cities. Ottawa samples consisted of primary sludge, whereas samples for all other tested locations were post-grit influent wastewater. This could potentially influence the PMMoV quantification and the corresponding calculated WC ratio because of a higher weight-normalized concentration of PMMoV found in solids issue primary sludge as compared to post-grit influent wastewater (data not shown). Additionally, other factors such as the intra-day variations of laboratory measurements, the inherent variations present due to the technology employed (PCR), and the various normalization techniques (or lack thereof) may also impact reported results. The measured viral signals (normalized with PMMoV for all locations in Ontario, non-normalized for locations in Alberta) are shown to have a relatively good visual agreement with clinical data throughout the studied periods (Figure 1a.). The COVID-19 pandemic in Canada has so far has been characterized by a surge of COVID-19 cases in the early pre-vaccination stage of the pandemic (March 2020) followed by four subsequent resurgences (Figure 1a.) in September 2020, December 2020, March 2021, and December 2021. The scale-standardized viral signal observed in wastewater at all seven tested sites demonstrates a good general agreement with the observed 7-day midpoint average for new COVID-19 clinical cases reported by their respective public health units over the separate resurgence periods, as determined via correlation analysis. A summary of the Pearson’s r correlation analyses is shown in the Supplementary Material (Table S3).

**Figure 1:**
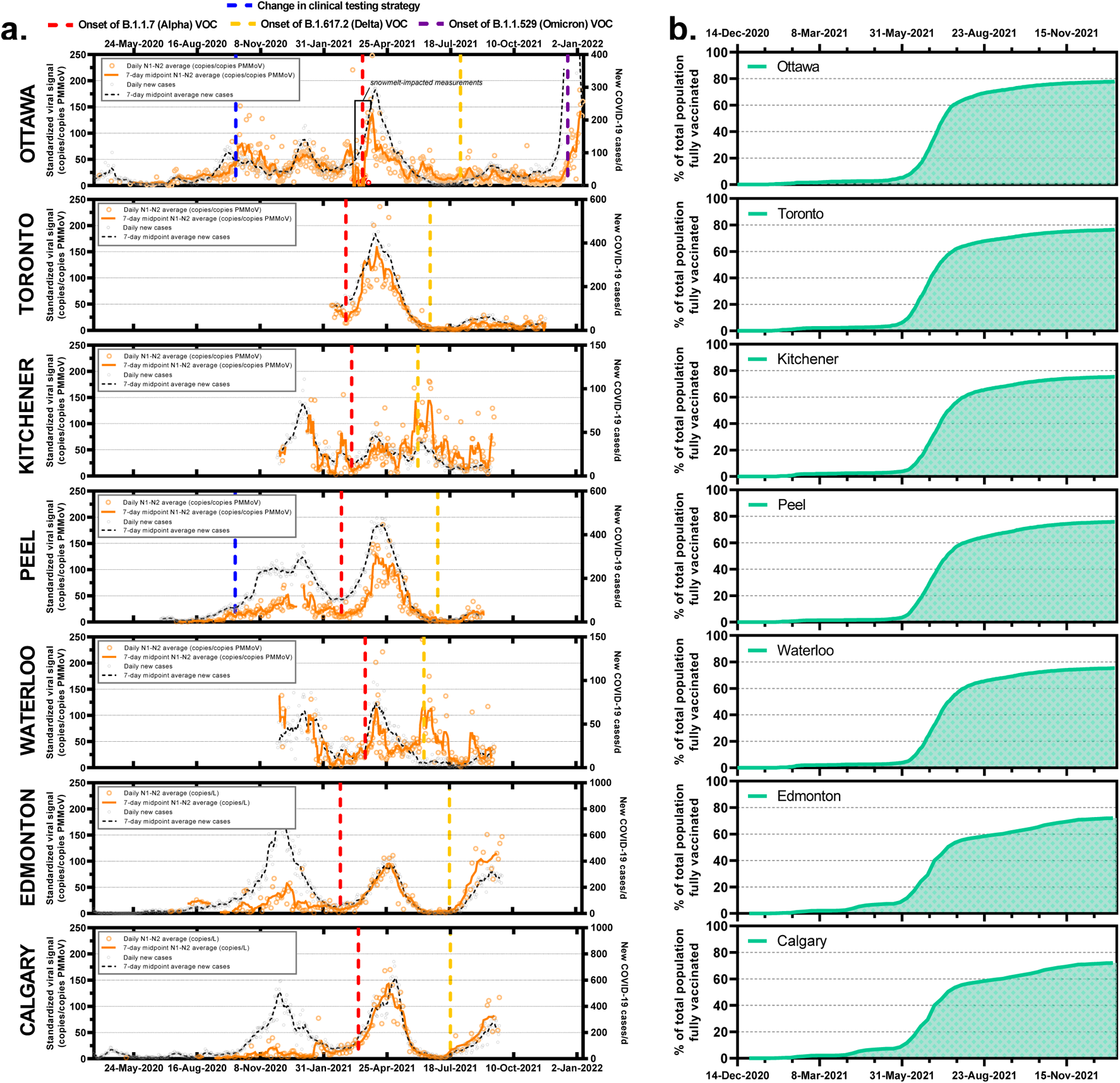
Comparison of a. scale-standardized PMMoV-normalized (Ottawa, Toronto, Kitchener, Peel and Waterloo) and non-PMMoV normalized (Edmonton and Calgary) SARS-CoV-2 viral signal measured in wastewater to reported new daily COVID-19 cases in the communities, with the onset of events of high epidemiological importance outlined with vertical dashed lines, coupled with b. summary of the immunization status of the populations living within the studied communities.

Mass immunization campaigns began in Canada in December 2020 for essential healthcare workers and the most vulnerable in the community (elderly and the immunocompromised). Vaccine rollout was relatively limited in early 2021, but vaccination rates increased rapidly throughout spring and summer. Facemask mandates were present throughout almost the entirety of the studied period, and intermittent gathering restrictions were also imposed intermittently. All studied communities reached a full immunization rate of at least 60% by Sept. 1st, 2021, and a full immunization rate (two approved vaccine doses) of at least 70% by Nov. 1st, 2021 (Figure 2b.).

### 3.2 The WC ratio as an indicator of key epidemiological occurrences in the cities

One of the proposed methods to relate wastewater to epidemiological information is the WC ratio, a ratio between wastewater viral titer and clinical cases was proposed by Xiao *et al*. (2022). As the ratio has the wastewater titers in the numerator and the reported new clinical cases in the denominator, Xiao et *al.* (2022) proposed that surveying the relative shifts in the WC ratio may yield information regarding clinical testing. They also reported that an increased WC ratio could be attributed to insufficient testing in the general population (i.e. testing not sufficient to capture the exponential growth of actual COVID-19 cases). When performing this particular analysis in all seven studied communities, significant changes in the WC ratio were hypothesized as being indicative of four major events: 1) a change in the clinical testing strategy that introduced the requirement to book a COVID-19 test appointment (5 out of 7 sites were in provinces that invoked this testing strategy); 2) the onset of the B.1.1.7 VOC during periods of low community immunity (all 7 sites were testing during this period), 3) the onset of the B.1.617.2 (Delta) VOC during periods of high community immunity (all 7 sites were testing during this period), and 4) the onset of the B.1.1.529 (Omicron) VOC during periods of high community immunization rates (75%+), with a VOC which has demonstrated immunity escape (a single site was testing during this period).

### 3.3 Event #1: WC ratio as an indicator of change in clinical testing strategy

Throughout the pandemic, public health units have adjusted their clinical testing strategies to better track the rate of incidence and prevalence of SARS-CoV-2 in their communities and help infer additional information, such as an approximate number of active disease cases and the pathogen’s reproduction rate (China Center for Disease Control, 2020; Hoseinpour Dehkordi et al., 2020; Hyafil and Moriña, 2020; Khailaie et al., 2021; Kolifarhood et al., 2020; Pan et al., 2020). PCR-based clinical testing methods have been the primary metric of COVID-19 rates of incidence and prevalence in communities for public health professionals. However, RT-PCR nucleic acid clinical tests are not without limitations; first, individuals may decline clinical testing or refuse to seek medical treatment during a pandemic despite being symptomatic, complicating both tracking of cases and contact tracing (Novoa et al., 2021), and second, RT-PCR test results may suffer from a substantial false-negative rate, especially when considering the time of testing in relation to symptom onset and the type of sample collected (sputum, stool, nasopharyngeal, oropharyngeal, etc.) (da Rocha Araujo, 2020; Hatchette, 2009; Wikramaratna et al., 2020).

An epidemiologically important event occurred when changes in clinical testing strategies were effected in the province of Ontario (affecting Ottawa, Kitchener, Toronto, Peel and Waterloo), Canada on October 6^th^, 2020. In response to an increasing backlog of tests, and to ensure individuals could get tested at a specific time with minimal wait (Crawlley et al., 2020), provincial guidelines and the framework for the initiation of SARS-CoV-2 RT-PCR clinical testing were changed from an open-access, walk-in format to one in which individuals were required to book an appointment before testing. This change in testing strategy presented a significant, if brief, barrier for Ontario residents wanting to be tested for COVID-19 by necessitating online bookings, which made it more difficult for groups of the general population to access testing resources. WWS had already been introduced in Ottawa and Peel during this period and, as such, a quantitative analysis of the effects of changes in testing strategies was performed.

When comparing the preceding 15-day period (Sept. 21^st^, 2020 – Oct. 5^th^, 2020) before the testing strategy change and the next 15-day period after enactment of the change (Oct. 6^th^, 2020 – Oct. 21^st^, 2020) in Ottawa the number of clinical tests performed on the general population decreased by an average of 23.2% (Figure 3a.). Concurrently, the local public health units reported a corresponding decrease of 16.8% in detected new clinical cases of COVID-19 in Ottawa and an increase of 22.1% in Peel (Figure 3a. and 3b.). During the same period, Ottawa’s normalized viral signal (average of N1-N2 viral copies/copies PMMoV) reported an increase of 55.9%, and the corresponding WC ratio increased by over 189% (Figure 3a.). Meanwhile, in Peel, the normalized viral signal increased by 19.7% increase while the WC ratio increased by only 7.5%. The important increase in WC ratio in Ottawa occurred due to both increased wastewater signal and decreasing numbers of reported new clinical cases occurring simultaneously, thereby amplifying the increase in the WC ratio. The weak increase in the WC ratio observed in Peel is hypothesized to have been caused by less frequent wastewater testing before and after the change in the testing strategy. The WWS program in Peel during this period was very close to inception and sampling was not as frequent before and after the change in the clinical testing strategy. During this period, variant testing in Ottawa’s wastewater confirmed that no known major VOC was circulating in the general population, and an analysis of the age demographic of new cases (Figure S1) showed no statistically significant difference during the 30-day sample set. Based on the observed rapid increase (>50%) in the WC ratio observed in Ottawa, it is hypothesized that the WC ratio may be an indicator of issues, such as flawed or difficult to access clinical testing resources, or overall insufficient clinical testing in relation to wastewater viral signal. This agrees with the findings and observations presented by Xiao et *al.* (2022).

**Figure 3:**
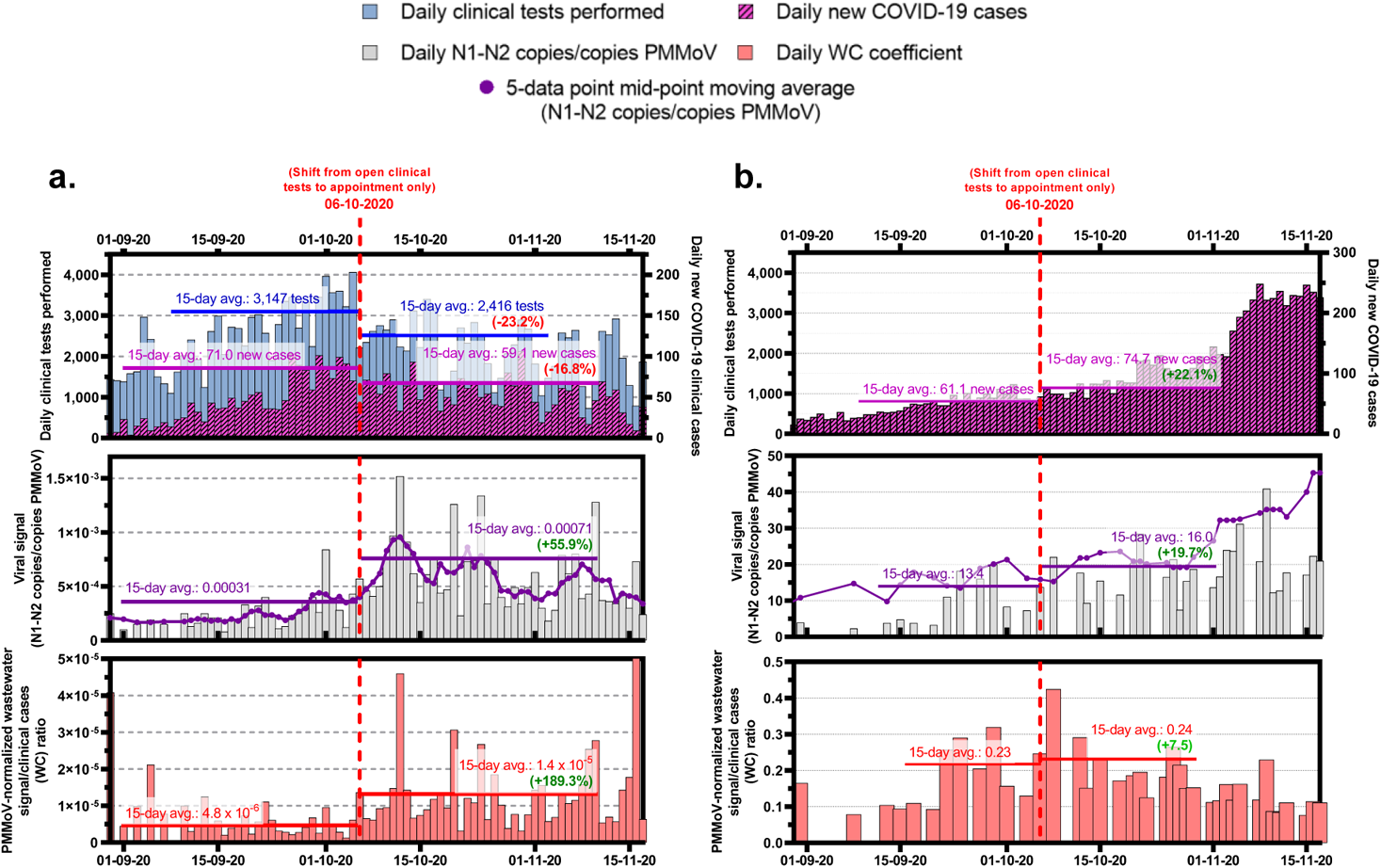
Comparison of the number of daily SARS-CoV-2 RT-PCR nucleic acid clinical tests performed and corresponding daily new COVID-19 positive cases, the daily viral signal measured through a SARS-CoV-2 wastewater surveillance program and the corresponding wastewater/clinical cases ratio (WC) during the same period, in a. Ottawa, and b. Peel.

While clinical testing currently remains the frontline measure of choice for the surveillance and tracking of COVID-19 incidence in the community, these findings strongly suggest that current and future disease surveillance efforts will benefit immensely from a multi-pronged surveillance approach incorporating more than the potentially biased case rates of clinically acquired diseases. COVID-19 and pandemic-potential diseases have been difficult to accurately track using traditional clinical testing methods as they are generally less effective at surveying disease burden due to many contributing factors, including testing capacity, the ability, and the desire of individuals to be tested (Gonsalves and Yamey, 2020; Jaiswal et al., 2020; Malinverni and Brigagão, 2020; Uscinski et al., 2020). It will continue to be difficult to conduct the widespread, mandatory disease surveillance of a population that is necessary to obtain information that could be interpreted as fully random and representative of the true rate of incidence of disease (McDermott and Newman, 2020; Pouwels et al., 2021; Rubin, 2020; Shao et al., 2020). In this context, it is therefore highly useful to utilize WWS and the WC ratio to complement traditional clinical testing results. WWS and the resulting measured viral load can be used as a confirmatory measure of actual rates of disease burden in the community, while rapid changes in the WC ratio may help identify periods of inadequate or altered clinical testing. Specifically, observing changes in the relative WC ratio may allow for the detection of changes in reported clinical cases that are not linked to actual changes in viral load in the community but are instead linked to shifts in testing rules and regulations, testing strategies, or fundamental changes in the disease vector. It is however important to also consider potential variations and uncertainties present in WWS (Ahmed et al., 2022; Li et al., 2021; Wade et al., 2022), as these will also have an impact on the WC ratio. It is hypothesized that in this study, due to trends observed during the change in testing strategies, is it likely that reported clinical cases may have been underreported in Ottawa and to a lesser extent, Peel, following the modification to the testing strategies applied in these communities. To determine the statistical significance of the change in WC ratio in both Ottawa and Peel during the studied periods, a non-parametric Mann-Whitney U test was performed, and the corresponding p-value was calculated. In Ottawa, the WC ratio before the change in strategy had a mean of 4.8 × 10^−6^ (IQR = 2.8 × 10^−6^ − 6.1 × 10^−6^), while the mean after the change in testing strategy was 1.4 × 10^−5^ (IQR = 8.2 × 10^−6^ − 1.4 × 10^−5^). As hypothesized, changes in the testing strategy correlated with an increase in WC ratio (p-value < 9 × 10^−5^). In Peel, the WC ratio before the change in testing strategy had a mean of 0.23 (IQR = 0.17 − 0.28), while the WC ratio after the change in testing strategy had a mean of 0.24 (IQR = 0.18 − 0.27). Change of the testing strategy in Peel did not appear to have a quantifiable effect on the WC ratio (p=0.886), however testing frequency was low during this period, which may explain the non-conclusive result.

#### 3.3.1 Event #2: WC ratio as an indicator of the onset of the B.1.1.7 (Alpha) variant of concern

SARS-CoV-2, being an RNA virus, is susceptible to mutations that can cause changes in its epidemiological characteristics, such as increased infectivity and/or virulence in comparison to its ancestral lineages (Burki, 2021; Drake and Holland, 1999; Duffy, 2018; Holmes and Rambaut, 2004). Throughout the COVID-19 pandemic, mutations of the SARS-CoV-2 spike (S) protein, referred to colloquially as D614G and N501Y, became hallmarks of the B.1.1.7 VOC, now referred to as the Alpha VOC. The onset of the Alpha VOC resulted in increased transmission of COVID-19 in the community (Frampton et al., 2021; Galloway et al., 2021; Graham et al., 2021), potentially affecting both observed viral load via WWS and new daily reported COVID-19 cases in the community by the public health unit. The period of onset of the Alpha VOC in the seven studied communities was confirmed via RT-qPCR allelic proportion determination (Figure 4) using a previously developed and validated assay (Graber et al., 2021). The onset of the Alpha VOC in a community was defined as such by determining when a consistent B.1.1.7 VOC allelic proportionality of between 40-60% of the SARS-CoV-2 viral signal in wastewater was reached and subsequently exceeded.

**Figure 4:**
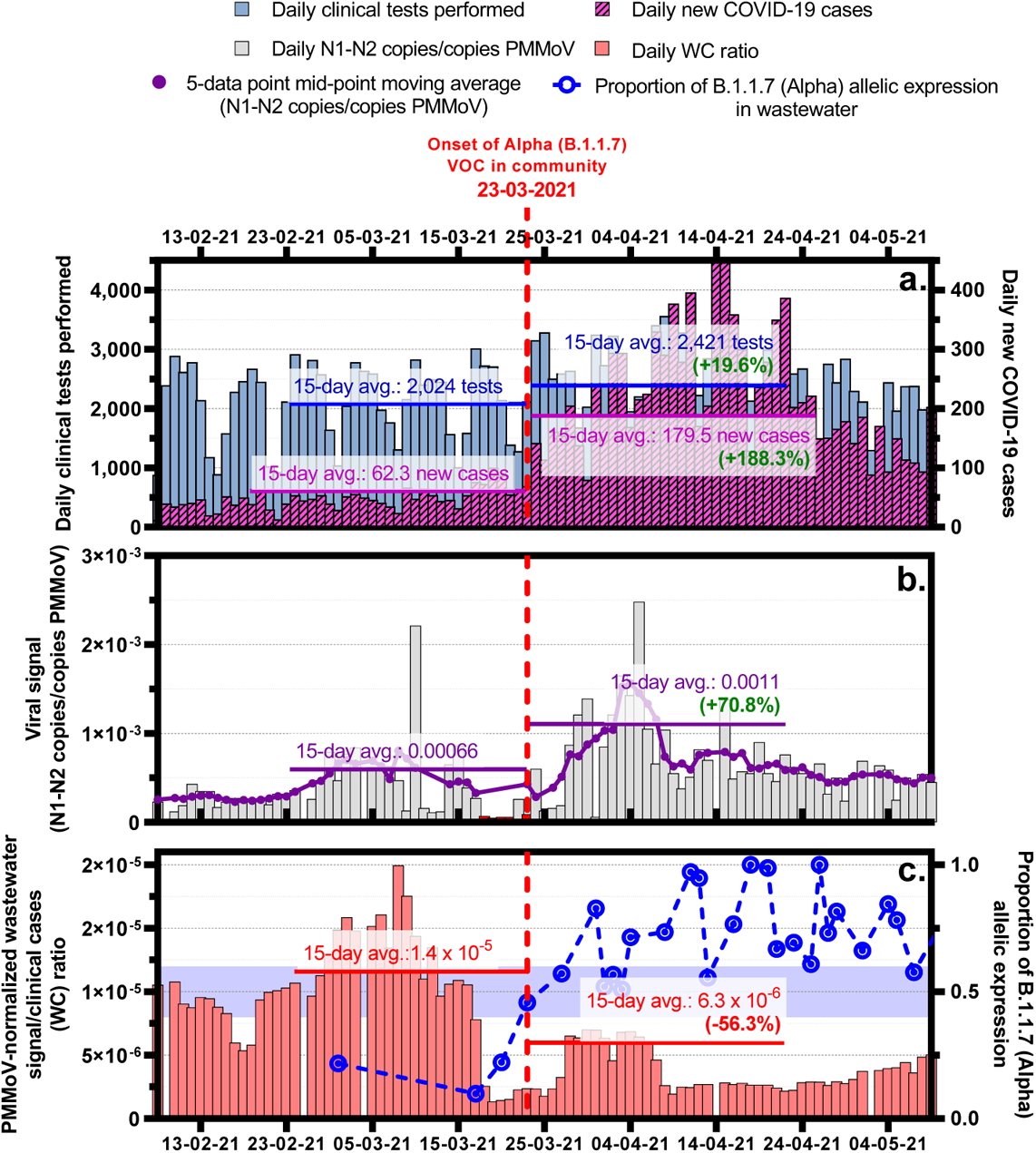
Comparison of a. the number of daily SARS-CoV-2 RT-PCR nucleic acid clinical tests performed and corresponding daily new COVID-19 positive cases in Ottawa, Canada, b. the daily viral signal measured through a SARS-CoV-2 wastewater surveillance program servicing approx. 91% of the City of Ottawa’s population (pop.: 1.1M) and c. the corresponding wastewater/clinical cases ratio (WC) during the same period, overlaid with the progression of the allelic expression of the B.1.1.7 (Alpha) VOC in wastewater.

Using Ottawa as an initial test case, when comparing the average of the preceding 15-day period and the average of the next 15-day period after the period of onset of the B.1.1.7 VOC, the number of clinical tests performed on the general population increased by 19.6% (Figure 4a.) and the local public health team reported a corresponding increase of 188.3% in the detection new daily clinical cases of COVID-19 (Figure 4b.). During the same period, normalized WWS reported a more modest increase of 70.8% in measured viral signal (average of N1-N2 viral copies/copies PMMoV). Furthermore, the WC ratio decreased by 56.3% (Figure 4c.). An analysis of the age demographic of new cases and changes in community immunization (Figure S2) showed no statistically significant difference during the studied 30-day period. The percentage of the total population fully immunized (2 doses +) increased from 2% to 2.3% during the studied 30-day period, but due to the time required for immuno-competence development and overall very low immunization rate, it is understood that vaccination was not likely to be an important factor in affecting reported clinical cases or viral fecal shedding dynamics during this period (Public Health Ontario, 2021).

A summary of proportional observed changes in the preceding 15-day period and following 15-day period in reported clinical cases of COVID-19, measured viral signal in wastewater and WC ratio at all seven tested locations is shown in the Supplementary Material (Table S4). Individually, proportional changes in reported clinical cases and measured SARS-CoV-2 viral signal in wastewater varied, however, in six of seven tested locations during this period, a marked decrease in the observed WC ratio (average decrease of 41.6%) was observed, signaling the emergence of the B.1.1.7 VOC as the dominant source of new infections (Figure 5). Only in Calgary did the WC ratio increase (6.4%) during the onset of the B.1.1.7 VOC. This trend in the decrease of the WC ratio signaling important chances in disease dynamics is consistent with observations by Xiao et *al.* (2022) for the Boston area during the period where the Alpha VOC became dominant in wastewater (Lee et al., 2021). The onset of the Alpha VOC led to an observed change in the relationship between the number of reported COVID-19 cases and the observed WWS viral signal, and its hypothesized that the Alpha VOC may have altered viral fecal shedding dynamics in terms of viral load in fecal matter over time (Miura et al., 2021). It has been suggested in the literature that there may be increased viral loading in symptomatic individuals caused by the Alpha variant as compared to the wild type of SARS-CoV-2 based on data obtained from nonhuman primates (Rosenke et al., 2021). However, there is not yet a consensus on this matter for human subjects (Badu et al., 2021; Zhu et al., 2021). The decrease in the WC ratio could also have been caused by a greater increase in transmissibility of the disease without a corresponding increase in fecal shedding (unequal variance in viral load in tissue and secretions), increased awareness in the general population, or increased severity of symptoms thereby leading more infected individuals to seek clinical testing, resulting into a lower WC ratio after the onset.

**Figure 5:**
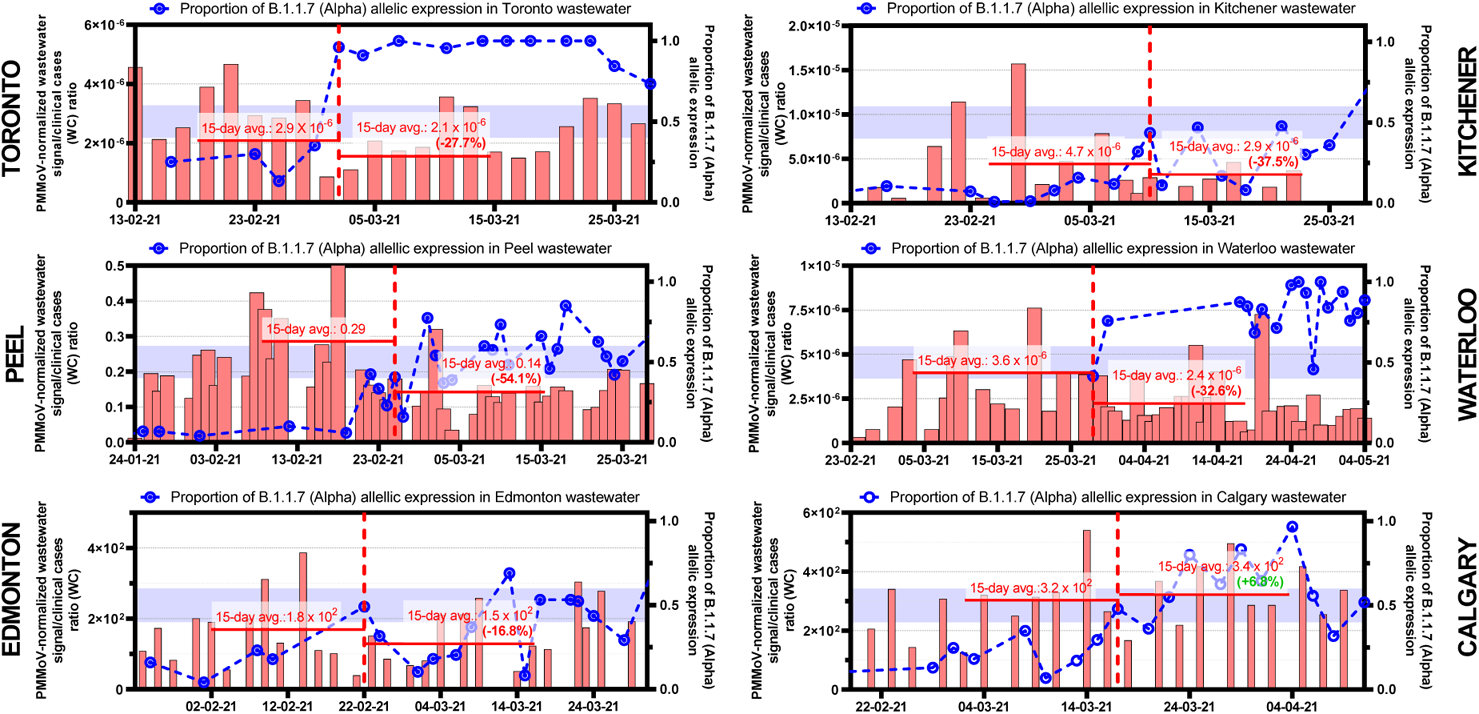
Comparison of the WC ratio and B.1.1.7 allelic proportionality during the onset of the B.1.1.7 (Alpha) VOC in the additional 6 locations in Canada (Toronto, Kitchener, Peel, Waterloo, Edmonton, and Calgary).

Mann-Whitney U tests were performed on each data set during the Alpha VOC onset to investigate the shifts in WC ratios in the studied locations. The results of the tests, along with accompanying interpretation, are shown below in Table 3.

**Table 3:** Mean WC ratio and interquartile ranges (1^st^ and 3^rd^ quartiles) before and after the Alpha VOC onset along with the results of Mann-Whitney U tests performed on the WC ratio data sets to evaluate the significance of shifts before and after the Alpha VOC onset. The resulting p-value, and textual interpretation are also shown to establish if the WC ratio shift was statistically significant.

| Location | Mean WC ratio<br>(before)/(after) | IQR<br>(1<sup>st</sup> / 3<sup>rd</sup> quartiles) | Mann-Whitney U test<br>p-value | Interpretation |
| --- | --- | --- | --- | --- |
| Ottawa | $1.4 \times 10^{-5}$ / $6.3 \times 10^{-6}$ | $4.5 \times 10^{-6} - 2.4 \times 10^{-5}$ / $4.1 \times 10^{-6} - 8.5 \times 10^{-6}$ | 0.254 | As hypothesized, decreases in the WC ratio occurred during the Alpha onset, however the Mann Whitney U test could not establish statistical significance ($p < 0.1$). |
| Toronto | $2.9 \times 10^{-6}$ / $2.1 \times 10^{-6}$ | $2.4 \times 10^{-6} - 3.6 \times 10^{-6}$ / $1.7 \times 10^{-6} - 2.7 \times 10^{-6}$ | 0.165 | As hypothesized, decreases in the WC ratio occurred during the Alpha onset however the Mann Whitney U test could not establish statistical significance ( $p < 0.1$ ). Lower frequency of testing could have had an impact on the test results. |
| Kitchener | $4.7 \times 10^{-6} / 2.9 \times 10^{-6}$ | $1.6 \times 10^{-6} - 6.3 \times 10^{-6} / 2.1 \times 10^{-6} - 3.5 \times 10^{-6}$ | 0.886 | As hypothesized, decreases in the WC ratio occurred during the Alpha onset, however the Mann Whitney U test could not establish statistical significance ($p < 0.1$). Lower frequency of testing could have had an impact on the test results. |
| Peel | 0.29 / 0.14 | 0.21 – 0.37 / 0.10 – 0.16 | 0.886 | As hypothesized, decreases in the WC ratio occurred during the Alpha onset, however the Mann Whitney U test could not establish statistical significance ($p < 0.1$). Lower frequency of testing could have had an impact on the test results. |
| Waterloo | $3.6 \times 10^{-6} / 2.4 \times 10^{-6}$ | $2.0 \times 10^{-6} - 3.7 \times 10^{-6} / 1.6 \times 10^{-6} - 2.9 \times 10^{-6}$ | 0.223 | As hypothesized, decreases in the WC ratio occurred during the Alpha onset, however the Mann Whitney U test could not establish statistical significance ($p < 0.1$). Lower frequency of testing could have had an impact on the test results. |
| Edmonton | $1.8 \times 10^2 / 1.5 \times 10^2$ | $1.1 \times 10^2 - 2.1 \times 10^2 / 1.9 \times 10^2 - 3.0 \times 10^2$ | 0.347 | As hypothesized, decreases in the WC ratio occurred during the Alpha onset, however the Mann Whitney U test could not establish statistical significance ($p < 0.1$). Lower frequency of testing or lack of normalization for flow/dilution effects could have had an impact on the test results. |
| Calgary | $3.2 \times 10^2 / 3.4 \times 10^2$ | $1.1 \times 10^2 - 2.1 \times 10^2 / 1.9 \times 10^2 - 3.0 \times 10^2$ | 0.391 | Contrary to hypothesized, increases in the WC ratio occurred during the Alpha onset, however the Mann Whitney U test could not establish statistical significance ($p < 0.1$). Lower frequency of testing or lack of normalization for flow/dilution effects could have had an impact on the test results. |

#### 3.3.2 Event #3: WC ratio as an indicator of the onset of the B.1.617.2 (Delta) variant of concern

Mutations in the SARS-CoV-2 genome at the N-terminal domain (NTD) and receptor-binding domain (RBD) locations led to the rise of the B.1.617 lineage first identified in October 2020 in India (Cherian et al., 2021; Yadav et al., 2021), culminating in the dominance of the Delta variant (B.1.617.2) in new COVID-19 infections in most locations in North America, including Canada. The Delta variant is characterized by increased transmissibility and virulence compared to the Alpha variant (Kannan et al., 2021; Liu et al., 2021; Planas et al., 2021). The periods of onset of the Delta VOC in the four studied communities with available data (Ottawa, Kitchener, Peel, and Waterloo) were confirmed via RT-qPCR allelic proportion determination of the D63G portion of the genome using an assay developed and validated in-house. These periods of onset were defined as reaching a B.1.617.2 VOC allelic proportion of 40-60% of the SARS-CoV-2 viral signal in wastewater and subsequently exceeding this proportionality.

Using Ottawa as an initial test case, when comparing the average of the preceding 15-day period and the average of the next 15-day period after the onset of the B.1.617.2 VOC a modest increase (14.0%) in clinical tests that were performed was observed (Figure 6a.). Meanwhile, during the same period, the reported new clinical cases of COVID-19 more than doubled, increasing by 121.1%. This trend was similar to what was observed during the onset of the Alpha variant for both the number of clinical tests performed and new daily COVID-19 cases in Ottawa. However, the total numbers of new reported COVID-19 cases per day were substantially lower by approximately one order of magnitude during the onset of the Delta variant. Specifically, a 15-day average of 9.1 new cases per day was observed during the onset of the Delta variant, while the 15-day average was 117.3 new cases per day during Alpha variant onset. These differences in observed new COVID-19 cases in the community were likely driven by immunization effects; as of Mar. 23^th^, 2021 (onset of Alpha variant), only 2.2% of the population in Ottawa was fully immunized, compared to 63.1% in Jul. 31^st^, 2021 (onset of the Delta variant) (Public Health Ontario, 2021). During the same period, the normalized WWS signal increased by 45.8% (average of N1-N2 viral copies/copies PMMoV), which is similar to the trend observed during the onset of the Alpha variant (44.5% increase). These combined factors led to a slight decrease of only 18.7% in the WC ratio during the onset of the Delta variant, compared to a significantly larger decrease of 56.3% during the Alpha variant onset. An analysis of the age demographic of new cases and changes in community immunization (Figure S3) showed no statistically significant difference during the studied 30-day period. It is hypothesized that immunization impact dampened both increases in reported new COVID-19 cases and viral signal in wastewater, effectively dampening the expected effects of the onset of a new, more infectious variant of SARS-CoV-2 on the WC ratio. Accordingly, communities with lower rates of immunization could be expected to exhibit greater reductions of the WC ratio due to increased rates of infections resulting in higher numbers of new daily COVID-19 cases. During the 30-day period where Delta emerged in Ottawa, full immunization in the total population (2 doses +) increased from 54.2% on Jul. 16^th^, 2021, to 66.9% on Aug. 14^th^, 2021), which likely limited the spread of the Delta variant within the community during this time, as attested by the observed viral load in wastewater and reported clinical cases (Figure 2a.).

**Figure 6:**
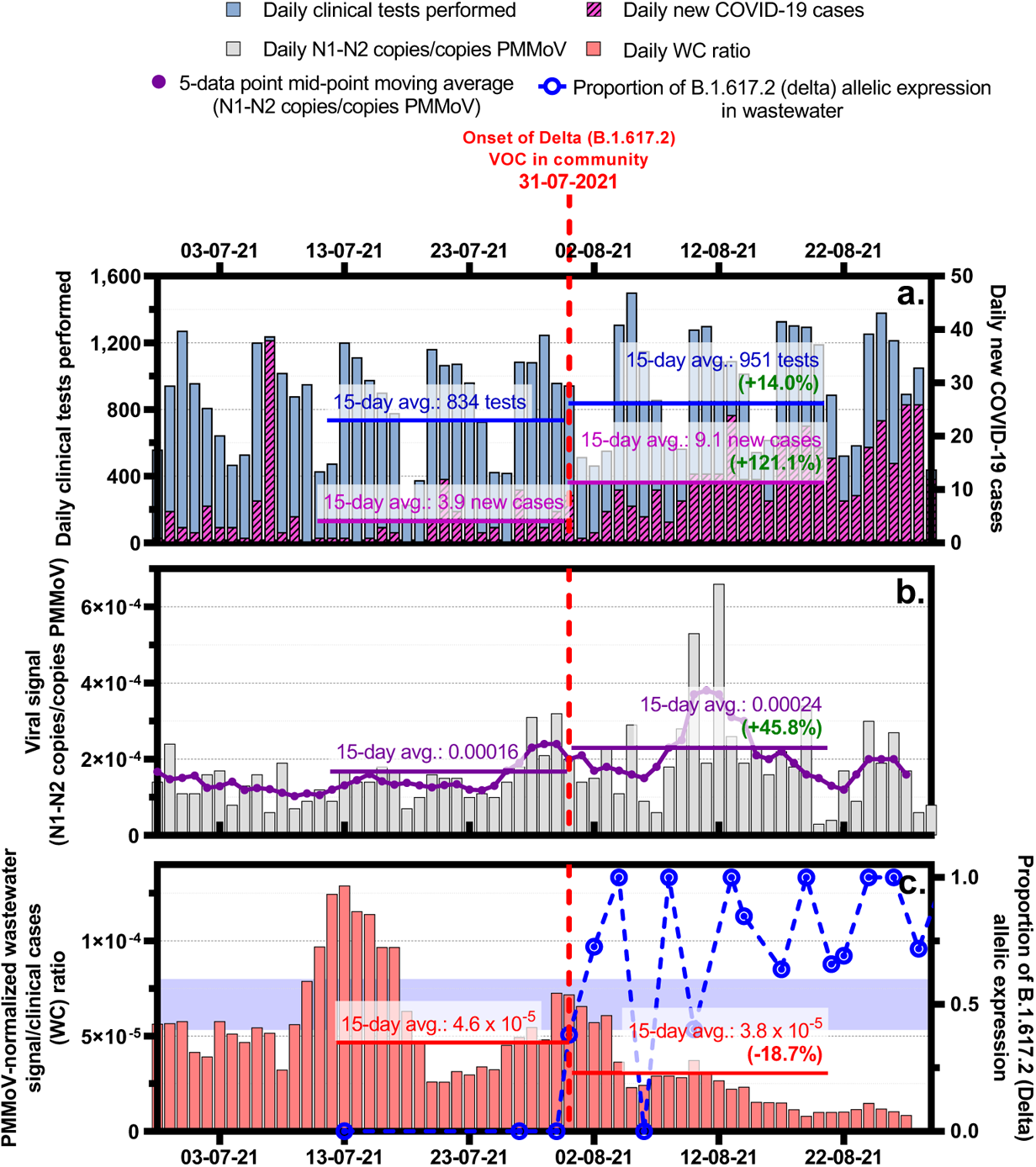
Comparison of a. the number of daily SARS-CoV-2 RT-PCR nucleic acid clinical tests performed and corresponding daily new COVID-19 positive cases in Ottawa, Canada, b. the daily viral signal measured through a SARS-CoV-2 wastewater surveillance program servicing approx. 91% of the City of Ottawa’s population (pop.: 1.1M) and c. the corresponding wastewater/clinical cases ratio (WC) during the same period, overlaid with the corresponding progression of the allelic expression of the B.1.617.2 (Delta) VOC in wastewater.

A summary of proportional observed changes in the preceding 15-day period and following 15-day period in reported clinical cases of COVID-19, measured viral signal in wastewater, and WC ratio at the tested locations is shown in the Supplementary Material (Table S4). Individually, proportional changes in reported clinical cases and measured SARS-CoV-2 viral signal in wastewater varied, however in three of the five tested locations (Ottawa, Toronto and Waterloo), the WC ratio decreased by an average of 27.0% during the emergence of the B.1.617.2 VOC as the dominant source of new infections. Meanwhile, in the four other tested locations with data during the emergence of the B.1.617.2 VOC (Peel, Kitchener, Edmonton and Calgary), the WC ratio instead increased by an average of 53.8%. A visual comparison of the WC ratio changes during the onset of the B.1.617.2 VOC is shown below in Figure. Contrary to trends observed during the B.1.1.7 VOC, the overwhelming trend in the WC ratio was not a decrease. It is believed that this increase in WC ratio in some of the locations (Peel, Kitchener, Edmonton and Calgary) may have been indicative of a disconnect between reported clinical cases in the communities and observed viral signal, caused partly by community immunization (hypothesized to lead to lower viral loads in feces) and local clinical testing capacity being reached or exceeded, leading to elevated WC ratio values caused by artificially low reported clinical cases. It is likely that in immunized communities without VOCs capable of immune escape, the change in the observed WC ratio will be lessened. Under such circumstances, the WC ratio is less indicative of the onset of new VOCs entering the community. Mann-Whitney U tests were performed on each data set during the Delta VOC onset to investigate the shifts in WC ratios in the studied locations. The results of the tests, along with accompanying interpretation, are shown below in Table 4.

**Figure 7:**
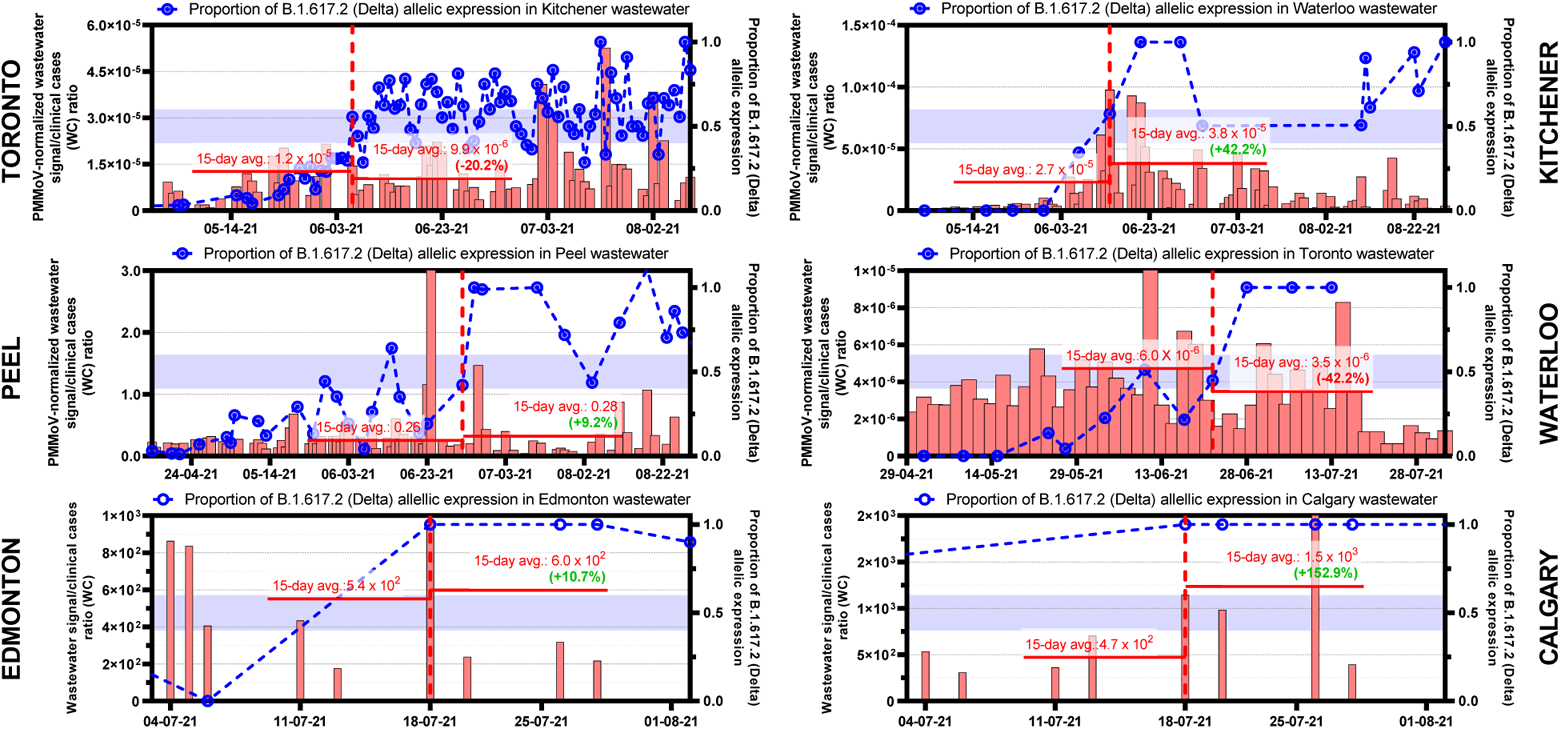
Comparison of the WC ratio and B.1.617.2 allelic proportionality during the onset of the B.1.617.2 (Delta) VOC in 6 additional locations in Canada (Toronto, Kitchener, Peel, Waterloo, Edmonton, and Calgary).

**Table 4:** Mean WC ratio and interquartile ranges (1^st^ and 3^rd^ quartiles) before and after the Delta VOC onset along with the results of Mann-Whitney U tests performed on the WC ratio data sets to evaluate the significance of shifts before and after the Delta VOC onset. The resulting p-value, and textual interpretation are also shown to establish if the WC ratio shift was statistically significant.

| Location | Mean WC ratio<br>(before)/(after) | IQR<br>(1<sup>st</sup> / 3<sup>rd</sup> quartiles) | Mann-Whitney<br>U test<br>p-value | Interpretation |
| --- | --- | --- | --- | --- |
| Ottawa | $4.6 \times 10^{-5}$ /<br>$3.8 \times 10^{-5}$ | $2.5 \times 10^{-5} - 6.4 \times 10^{-5}$ /<br>$1.5 \times 10^{-5} - 4.3 \times 10^{-5}$ | 0.079 | As hypothesized, decreases in the WC ratio occurred during the Delta onset, and the Mann Whitney U test could establish statistical significance ($p < 0.1$). |
| Toronto | $6.0 \times 10^{-6}$ /<br>$3.5 \times 10^{-6}$ | $3.0 \times 10^{-6} - 6.4 \times 10^{-6}$ /<br>$2.1 \times 10^{-6} - 3.5 \times 10^{-6}$ | 0.132 | As hypothesized, decreases in the WC ratio occurred during the Delta onset, however the Mann Whitney U test could not establish statistical significance ($p < 0.1$). |
| Kitchener | $1.2 \times 10^{-5}$ /<br>$9.9 \times 10^{-6}$ | $8.1 \times 10^{-6} - 1.8 \times 10^{-5}$ /<br>$7.3 \times 10^{-6} - 1.1 \times 10^{-5}$ | 0.409 | As hypothesized, decreases in the WC ratio occurred during the Delta onset, however the Mann Whitney U test could not establish statistical significance ($p < 0.1$). Lower frequency of testing could have had an impact on the test results. |
| Peel | 0.26 /<br>0.28 | 0.16 – 0.32 /<br>0.24 – 0.45 | 0.257 | Contrary to the emitted hypothesis, increases in the WC ratio occurred during the Delta onset, however the Mann Whitney U test could not establish statistical significance ($p < 0.1$). Lower frequency of testing could have had an impact on the test results. |
| Waterloo | $2.7 \times 10^{-5}$ /<br>$3.8 \times 10^{-5}$ | $6.1 \times 10^{-6} - 2.7 \times 10^{-5}$ /<br>$2.2 \times 10^{-5} - 6.1 \times 10^{-5}$ | 0.065 | Contrary to the emitted hypothesis, increases in the WC ratio occurred during the Delta onset, and the Mann Whitney U test established statistical significance ( $p < 0.1$ ). Lower frequency of testing could have had an impact on the test results. |
| Edmonton | $5.4 \times 10^2 / 6.0 \times 10^2$ | $4.1 \times 10^2 - 8.4 \times 10^2 / 2.6 \times 10^2 - 9.0 \times 10^2$ | 0.201 | Contrary to hypothesized, increases in the WC ratio occurred during the Delta onset, however the Mann Whitney U test could not establish statistical significance ($p < 0.1$). Lower frequency of testing or lack of normalization for flow/dilution effects could have had an impact on the test results. |
| Calgary | $4.7 \times 10^2 / 1.5 \times 10^3$ | $3.5 \times 10^2 - 5.8 \times 10^2 / 7.5 \times 10^2 - 1.1 \times 10^3$ | 0.624 | Contrary to hypothesized, increases in the WC ratio occurred during the Delta onset, however the Mann Whitney U test could not establish statistical significance ($p < 0.1$). Lower frequency of testing or lack of normalization for flow/dilution effects could have had an impact on the test results. |

#### 3.3.3 Event #4: WC ratio as an indicator of the onset of the B.1.1.529 (Omicron) variant of concern

Later in the pandemic, further mutations on the S protein RBD locations resulted in a very genotypically different VOC, denoted B.1.1.529 (Omicron). The combination of several mutations observed previously in the C.37 (Lambda) variant and several new mutations on the S protein RBD resulted in a significantly more infectious yet symptomatically milder VOC (Halfmann et al., 2022; Shuai et al., 2022), which spread rapidly around the world and replacing Delta as the main VOC in the majority of the location where it established. One of the important characteristics of Omicron is its ability to potentially evade neutralization in sera, even in vaccine-immunized individuals (He et al., 2021; Zhang et al., 2021).

Using Ottawa as an initial test case, when comparing the average of the preceding 15-day period and the average of the next 15-day period after the period of onset of the B.1.1.529 VOC (December 20^th^, 2021), the number of clinical tests performed on the general population increased by 29.6% and the local public health reported a corresponding increase of 497.3% in the detection new clinical cases of COVID-19 (Figure 8a.). During the same period, normalized WWS reported an important increase of 243.5% in measured viral signal (average of N1-N2 viral copies/copies PMMoV). At the same time, the WC ratio decreased by 41.9% (Figure 8c.). It is believed that significant decreases in the WC ratio are an indicator of the onset of new VOCs with immune escape abilities. An analysis of the age demographic of new cases and changes in community immunization (Figure S4) showed no statistically significant difference during the studied 30-day period. Daily vaccination rates during the studied 30-day period increased slightly over time due to residents obtaining third-dose booster vaccinations, but due to the time required for immuno-competence development, it is understood that vaccination was not likely to be a major factor influencing changes of the WC ratio.

**Figure 8:**
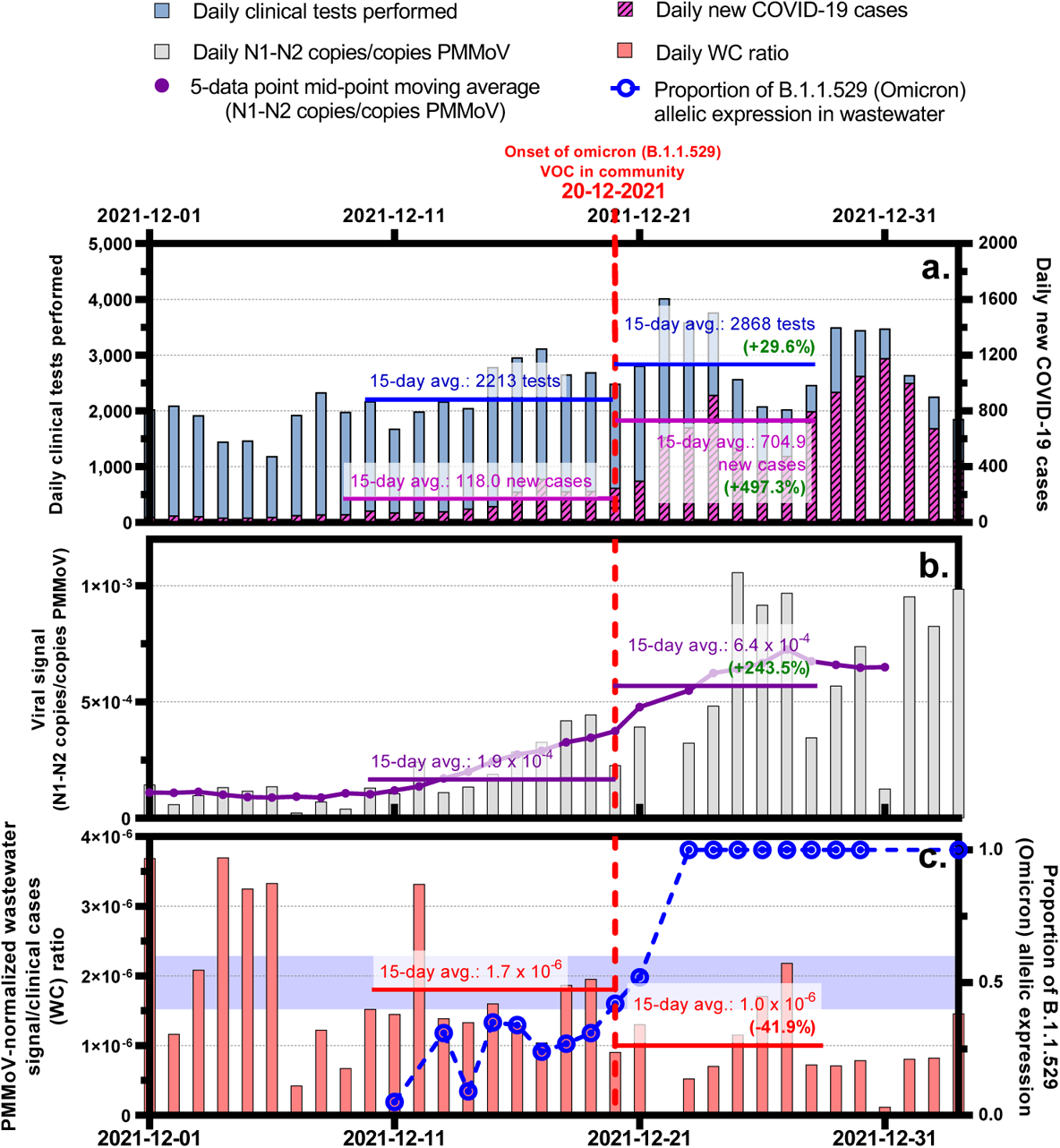
Comparison of a. the number of daily SARS-CoV-2 RT-PCR nucleic acid clinical tests performed and corresponding daily new COVID-19 positive cases in Ottawa, Canada, b. the daily viral signal measured through a SARS-CoV-2 wastewater surveillance program servicing approx. 91% of the City of Ottawa’s population (pop.: 1.1M) and c. the corresponding wastewater/clinical cases ratio (WC) during the same period, overlaid with the corresponding progression of the allelic expression of the B.1.1.529 (Omicron) VOC in wastewater.

### 3.4 Discussion

The in-depth analysis of the WC ratio during several major epidemiological events (as described in the above sections) revealed several major trends and findings: First, performed alone, both traditional clinical testing and WWS sometimes provide limited information for public health units that are monitoring a major pandemic disease like COVID-19. Clinical testing appears to sometimes suffer from underreporting due to the limited reach of the clinical tests, coupled with the limits in the ability and the desire of individuals to be tested, especially if they do not feel unhealthy. Furthermore, a strong relationship exists between the number of clinical tests performed and the number of detected new cases in the community, particularly during periods of disease resurgence, where a high degree of community transmission occurs, potentially biasing interpretations of results. It is hypothesized that this weakness in traditional clinical testing is due to the difficulty in performing enough tests throughout the community to constitute testing rates that approximate a random distribution. In contrast, WWS effectively tests all individuals connected to the sewershed regardless of whether they are symptomatic or believe they have been exposed to the virus. Monitoring the WC ratio while performing RT-qPCR-based WWS can accurately detect the time of arrival of new variants of concern in a community during a pandemic as quickly as traditional clinical testing and does not require infected individuals to seek clinical testing or enter the healthcare system. Throughout the study, it appears that a shift in the rate of change of the WC ratio (slowing of a decreasing trend) and the relationship between reported clinical cases and observed WWS signal occurred for three reasons: (i) immunization may significantly reduce the number of individuals seeking COVID-19 testing due to a reduction of apparent symptoms caused by vaccination; (ii) as the pandemic progresses, disinterest may develop in the population, leading to a lower likelihood of symptomatic individuals seeking clinical testing, particularly if they have been immunized; and (iii) immunization may affect individual fecal shedding characteristics, impacting observed viral signal in wastewater. However, at present, it is still uncertain if this is the case, although preliminary results and unpublished data suggest this trend may exist (Bivins and Bibby, 2021).

Finally, during the onset of the highly infectious and immunity evading Omicron variant, very strong increases in reported clinical cases and observed viral signal in wastewater co-occurred with a strong decrease in the WC ratio. It is hypothesized that the magnitude of change in the WC ratio in a longitudinal analysis of a single testing location may correlate with the overall disease burden in the community (Figure 9). At points of inflection in the WC ratio, caused by events of major epidemiological importance, such as the onset of epidemiologically relevant variants, the rate of change appears to correlate with the disease burden. As an example, the direction and rate of change in the WC ratio during the onset of the Alpha and Omicron variants were similar and distinctly different from the period of onset of the Delta variant in Ottawa. An apparent distinction between the onset of Alpha and Delta variants also exists in the studied communities with the required data (% allelic proportions calculated for Alpha and Delta variants). To determine the statistical significance of the change in WC ratio in Ottawa during the Omicron VOC onset, a non-parametric Mann-Whitney U test was performed, and the corresponding p-value was calculated. The WC ratio before the Omicron VOC onset had a mean of 1.7 × 10^−6^ (IQR = 1.2 × 10^−6^ – 1.8 × 10^−6^), while the mean after the Omicron VOC onset was of 1.0 × 10^−6^ (IQR = 7.2 × 10^−7^ − 1.3 × 10^−6^). As hypothesized, the onset of a more infectious VOC (Omicron) corresponded with a decrease in WC ratio (p-value < 0.035).

**Figure 9:**
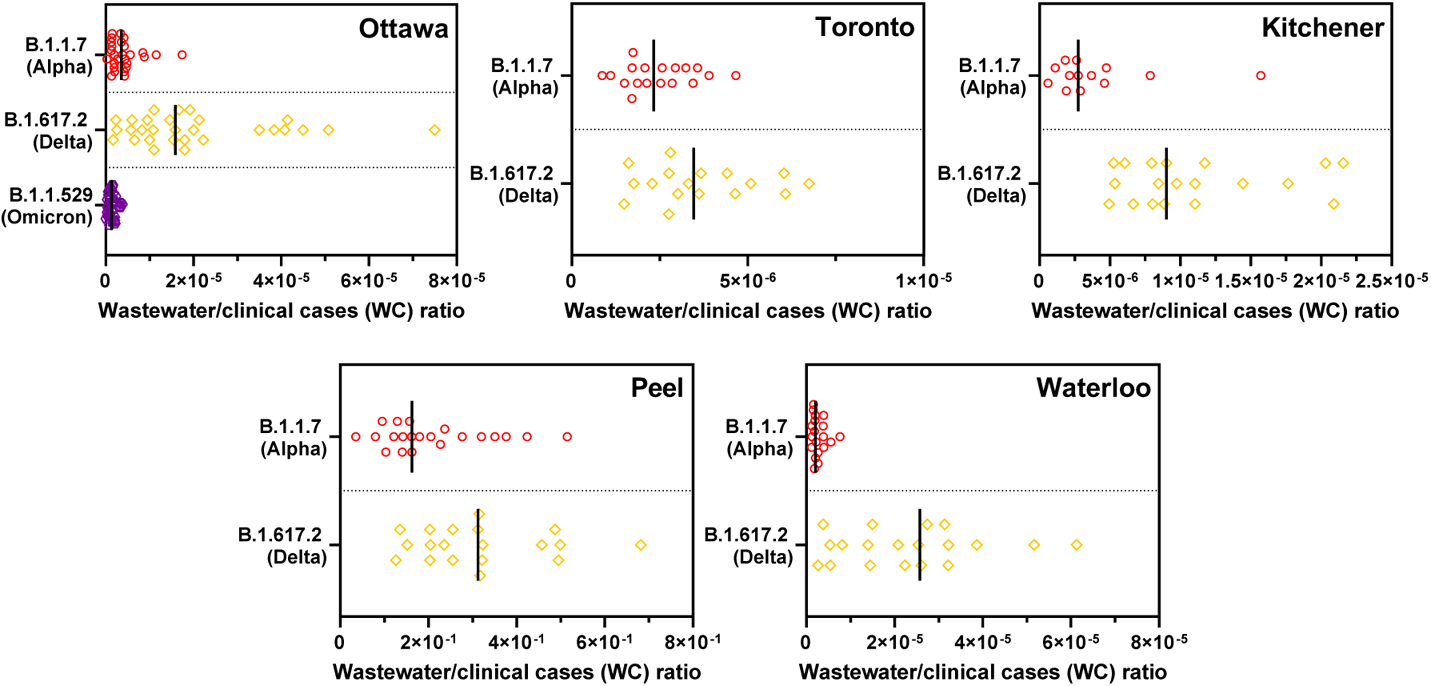
Comparison of WC ratio amplitude during the 30-day periods of onset of the B.1.1.7 (Alpha), B.1.617.2 (Delta) and B.1.1.529 (Omicron) VOCs in Ottawa, Kitchener, Peel and Waterloo.

It is important that in the interpretation of shifts in the WC ratio, relative changes are to be investigated, rather than absolute changes, due to the site-specific nature of the WC ratio. Slight modifications to methodologies and techniques in the laboratory, as well as molecular targets analyzed, could cause differences in measurements, and therefore change the magnitude of the WC significantly. This is observed in this study as the different laboratories employ similar methods, but intricate differences regarding sample processing volumes and sample solids mass appear to significantly affect results and the feasibility of direct comparisons. Furthermore, it is important to also acknowledge that in this study, 5 of the 7 studied communities (Ottawa, Toronto, Kitchener, Peel, and Waterloo) were in one province (Ontario), while the 2 other communities (Edmonton and Calgary) were situated in another (Alberta), which may have led to slight differences in both employed epidemic control measures and access to PCR testing, which could also influence the WC ratio. There still exists a gap of knowledge in normalizing the WC ratio before it could be used to compare data sets from completely different communities, however, on a single community level, or in instances where a single, unified methodology and analysis is applied, comparisons of the WC ratio between communities should also be possible.

## 4 Conclusions

This study strongly suggests that regular, daily monitoring of the WC ratio can reveal and detect the onset of changes in disease transmission patterns, and the arrival/onset and waning of more infectious variants or mutations of a pathogen or disease. When using traditional clinical case data and WWS, additional monitoring of the WC ratio may provide a greater understanding of the surveillance metrics during events of high epidemiological importance. The specific conclusions of this study are as follows: During periods of resurgence or increases in disease spread in a community, monitoring of reported clinical cases may provide a limited understanding of the true disease prevalence rates when community daily clinical testing is restricted or insufficient. The WC ratio will strongly increase when the disease burden increases in a community where an insufficient number of clinical tests are being performed to adequately approximate the disease burden in the community. Furthermore, if a community is not well immunized (less than 2.5% of individuals have received two doses of a COVID-19 vaccine), based on observed trends in this study, the onset of new, more infectious VOCs may lead to a decrease in the WC ratio. Meanwhile, if a community is well immunized (>60.0% of residents have received two doses of a COVID-19 vaccine), the onset of new, more infectious VOCs caused modest decreases in the WC ratio. Moreover, the onset of the Omicron VOC resulted in significant decreases in the WC ratio, as was observed during Alpha VOC dominance in early 2021, and this, irrespective of the immunization status of the community, is likely due to the immune escape capability of the Omicron VOC. It was observed that as a result, decreases in the WC can often be associated with the onset of a change in disease dynamics and the arrival of more infectious novel variants in the community and could be used as a tool to identify the emergence of new variants in the future. Finally, it was determined that changes in the magnitude and inflection points in the WC ratio at a longitudinally surveyed location may be indicative of changes in disease burden and the ability of the pathogen to spread. These findings indicate that monitoring the WC ratio during the SARS-CoV-2 pandemic and other, future pandemics, will yield additional insight into the disease burden in a community and should be performed where possible.

## Data Availability

All data produced in the present study are available upon reasonable request to the authors

## Acknowledgements

The authors wish to acknowledge the help and assistance of the University of Ottawa, the University of Toronto, the University of Waterloo, and the University of Alberta for their important contributions to the laboratories performing wastewater analysis. Furthermore, authors are very thankful of the support and continued cooperation of all wastewater treatment facility operators (City of Ottawa, the City of Toronto, the City of Waterloo, the Region of Peel, the City of Kitchener, EPCOR Utilities and the City of Calgary). Additional thanks go out to the Ottawa Hospital, the Children’s Hospital of Eastern Ontario, the Children’s Hospital of Eastern Ontario’s Research Institute, Ottawa Public Health, Toronto Public Health, Peel Public Health, the Region of Waterloo Public Health and Emergency Services, Public Health Ontario and all their employees involved in the project during this study. Their time, facilities, resources, and thoughts provided throughout the study helped the authors greatly.

