## Supplemental Files for "Wastewater to clinical case (WC) ratio of COVID-19 identifies insufficient clinical testing, onset of new variants of concern and population immunity in urban communities"

### Supplemental Material

Table S1: Summary of studied water resource recovery facilities' treatment characteristics.

<table>
<tr><th>Water resource recovery facility</th><th>Average daily flow (MLD)</th><th>Treatment level (Primary, Secondary, Tertiary)</th><th>Approx. population deserved</th></tr>
<tr><td>Ottawa (Robert O. Pickard Environmental Centre)</td><td>435</td><td>Secondary</td><td>910,000</td></tr>
<tr><td>Toronto (Ashridge’s Bay water resource recovery facility)</td><td>556</td><td>Secondary</td><td>1,603,700</td></tr>
<tr><td>Toronto (Highland Creek water resource recovery facility)</td><td>173</td><td>Secondary</td><td>533,000</td></tr>
<tr><td>Toronto (North Toronto water resource recovery facility)</td><td>46</td><td>Secondary</td><td>55,000</td></tr>
<tr><td>Kitchener (Kitchener water resource recovery facility)</td><td>70</td><td>Secondary</td><td>262,300</td></tr>
<tr><td>Peel (Clarkson water resource recovery facility)</td><td>446</td><td>Secondary</td><td rowspan="2">1,500,000</td></tr>
<tr><td>Peel (G.E. Booth water resource recovery facility)</td><td>236</td><td>Secondary</td></tr>
<tr><td>Waterloo (Waterloo water resource recovery facility)</td><td>46</td><td>Secondary</td><td>162,000</td></tr>
<tr><td>Edmonton (EPCOR Gold Bar water resource recovery facility)</td><td>285</td><td>Tertiary</td><td>834,000</td></tr>
<tr><td>Calgary water resource recovery facility</td><td>420</td><td>Tertiary</td><td>1,000,000</td></tr>
</table>

Table S2: Oligonucleotide sequences used for the detection of targets in this study.

| Primer/probe & supplier | Sequence | Reference |
|---|---|---|
| 2019-nCoV_N1 forward primer (IDT) | GAC CCC AAA ATC AGC GAA AT | [1] |
| 2019-nCoV_N1 reverse primer | TCT GGT TAC TGC CAG TTG AAT CTG | [1] |
| 2019-nCoV_N1 probe (IDT) | **6-FAM**-ACC CCG CAT/**ZEN**/ TAC GTT TGG TGG ACC-**IBFQ** | [1] |
| 2019-nCoV_N2 forward primer | TTA CAA ACA TTG GCC GCA AA | [1] |
| 2019-nCoV_N2 reverse primer | GCG CGA CAT TCC GAA GAA | [1] |
| 2019-nCoV_N2 probe (IDT) | **6-FAM**-ACA ATT TGC/**ZEN**/CCC CAG CGC TTC AG-**IBFQ** | [1] |
| PMMoV forward primer | GAG TGG TTT GAC CTT AAC GTT GA | [2] |
| PMMoV reverse primer | TTG TCG GTT GCA ATG CAA GT | [2] |
| PMMoV probe (ABI) | **6-FAM**-CCT ACC GAA GCA AAT G-**MGB** | [2] |
| VSV forward primer | ATA AGA TAC CGG GCT TGC AC | [3] |
| VSV reverse primer | ACA AAG ACA TGC CCG ACA C | [3] |
| VSV probe (ABI) | **6-FAM**-CCA TGT TGT ATT TGG ACC C-**MGB** | [3] |
| D3 (non-B.1.1.7) forward primer | ATC TAA ACG AAC AAA CTA AAA TGT CTG AT | [4] |
| D3L (B.1.1.7) forward primer | CAT CTA AAC GAA CAA ACT AAA TGT CTC TA | [4] |
| D63G (B.1.617.2) forward primer (ABI) | TCA CTC AAC ATG GCA AGA AAG G | *Personal communication with M. Fuzzen* |
| D63G (B.1.617.2) reverse primer (ABI) | GGT AGT AGC CAA TTT GGT CAT CT | *Personal communication with M. Fuzzen* |
| N63 (non-B.1.617.2) forward primer | CTC ACT CAA CAT GGC AAG AAA G | *Personal communication with M. Fuzzen* |
| D63G probe | **6-FAM**-CCT TAA ATT CCC TCG ATG ACA AGG CG-**MGB** | *Personal communication with M. Fuzzen* |
| N200 (B.1.617.2) forward primer | TAG TCG CAA CAG TTC AAG AAA T | [5] |
| N200 (B.1.617.2) reverse primer | CTG GTT CAA TCT GTC AAG CAG | [5] |
| N200 (B.1.617.2) universal probe | **6-FAM-**TCC TGC TAG AAT GGC-**BHQ-1** | [5] |
| N200 (B.1.617.2) delta probe | **CAL Fluor Orange 560**-CAG CAG TAT GGG AAC T-**BHQ-2** | [5] |
| C28311 (P13L) (non-B.1.1.529) forward primer | CCA AAA TCA GCG AAA TGA ACC | *This study* |
| C28311T (P13L) (B.1.1.529) forward primer | CCA AAA TCA GCG AAA TGA ACT | *This study* |
| N1 (version 2) probe (ABI) | **6-FAM**-CCG CAT TAC GTT TGG TGG ACC C-**MGB** | *This study* |

Table S3: Summary of regression analyses during different resurgences of the COVID-19 pandemic in seven Canadian cities, showing good to excellent correlations between daily observed clinical COVID-19 cases and measured normalized SARS-CoV-2 viral signal in wastewater.

| Location | Pearson's r, p-value, n; during 1st resurgence | Pearson's r, p-value, n; during 2nd resurgence | Pearson's r, p-value, n; during 3rd resurgence | Pearson's r, p-value, n; during 4th resurgence |
|---|---|---|---|---|
| **Ottawa** | R=0.263, p-value = 0.01, n = 91 | R = 0.878, p-value < 0.01, n = 59 | R = 0.760, p-value < 0.01, n = 90 | R = 0.446, p-value = 0.02, n = 27 |
| **Toronto** | - | - | R = 0.935, p-value < 0.01, n = 68 | - |
| **Kitchener** | - | R = 0.831, p-value < 0.01, n = 32 | R = 0.383, p-value < 0.01, n = 61 | - |
| **Peel** | R = 0.856, p-value < 0.01, n = 124 | R = 0.686, p-value < 0.01 n = 53 | R = 0.910, p-value < 0.01, n = 131 | - |
| **Waterloo** | - | - | R = 0.737, p-value < 0.01, n = 141 | - |
| **Edmonton** | - | R = 0.012, p-value = 0.895, n = 62 | R = 0.985, p-value < 0.01, n = 56 | - |
| **Calgary** | - | R = 0.845, p-value < 0 .01, n = 69 | R = 0.973, p-value < 0.01, n = 45 | - |

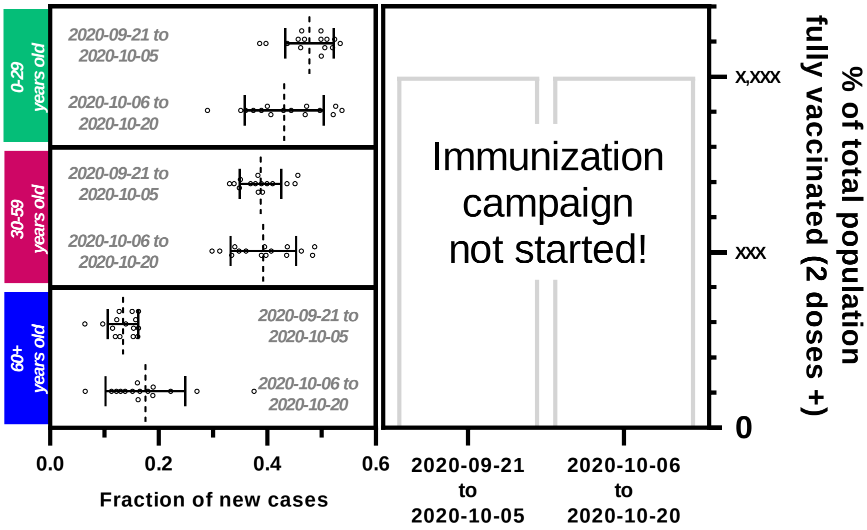


Figure S1: Comparison of the demographic distribution of new cases, per age group, and the proportion of the total population which was fully immunized (2 doses +) during the 15-day period preceding and following the change in testing rules and regulations on Oct. 6th, 2020, in Ottawa, Canada.

Table S4: Proportional changes in reported clinical cases of COVID-19, measured WWS SARS-CoV-2 viral signal and WC ratio at all seven tested locations between the preceding 15-day and following 15-day periods where 40-60% allelic proportionality of the Alpha (B.1.1.7) VOC was measured in wastewater.

| **Location** | **Date where allelic proportionality of B.1.1.7 VOC reached 40-60%** | **Change in reported clinical cases (%)** | **Change in measured SARS-CoV-2 viral signal in wastewater (%)** | **Change in WC ratio (%)** |
|---|---|---|---|---|
| Ottawa | 23-03-2021 | +188.3 | +70.8 | -56.3 |
| Toronto | 02-03-2021 | +45.4 | +9.2% | -27.7 |
| Kitchener | 10-03-2021 | -20.9 | -65.7 | -37.5 |
| Peel | 22-02-2021 | +9.7 | +2.3 | -54.1 |
| Waterloo | 27-03-2021 | +224.0 | +119.3 | -32.6 |
| Edmonton | 23-02-2021 | +24.3 | -10.8 | -16.8 |
| Calgary | 22-03-2021 | +82.7 | +92.5 | +6.4% |

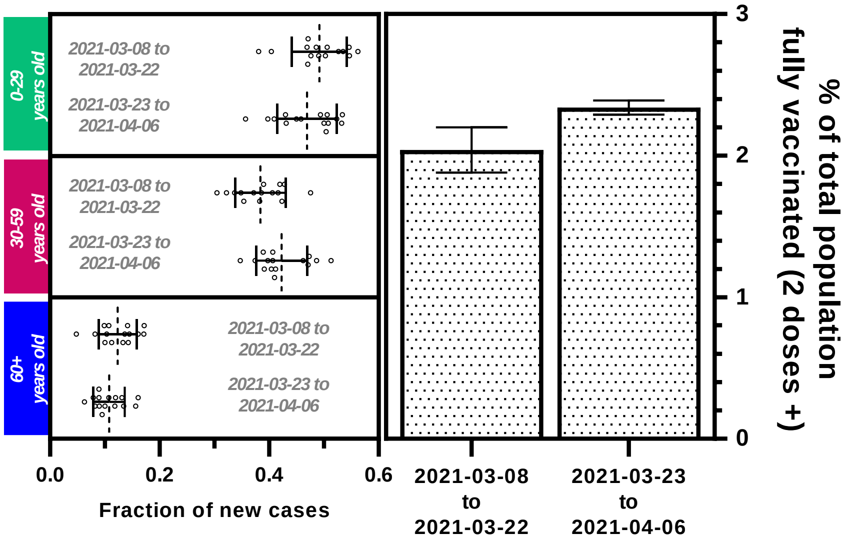


Figure S2: Comparison of the demographic distribution of new cases, per age group, and the proportion of the total population which was fully immunized (2 doses +) during the 15-day period preceding and following the onset of the Alpha (B.1.1.7) variant of concern in Ottawa, Canada.

38

Table S5: Proportional changes in reported clinical cases of COVID-19, measured WWS SARS-CoV-2 viral signal and WC ratio at all 4 tested locations with data between the preceding 15-day and following 15-day periods where 40-60% allelic proportionality of the Delta (B.1.617.2) VOC was measured in wastewater.

| **Location** | **Date where allelic proportionality of B.1.617.2 VOC reached 40-60%** | **Change in reported clinical cases (%)** | **Change in measured SARS-CoV-2 viral signal in wastewater (%)** | **Change in WC ratio (%)** |
|---|---|---|---|---|
| Ottawa | 31-07-2021 | +121.1 | +45.8 | -18.7 |
| Toronto | 22-06-2021 | -53.5 | -75.1 | -42.2 |
| Kitchener | 06-06-2021 | +51.7 | +29.1 | -20.2 |
| Peel | 28-05-2021 | -63.3 | -52.3 | +9.2 |
| Waterloo | 02-07-2021 | -26.8 | +17.9 | +42.2 |
| Edmonton | 18-07-2021 | +168.0 | +210.9 | +10.7 |
| Calgary | 18-07-2021 | +185.7 | +533.8 | +152.9 |

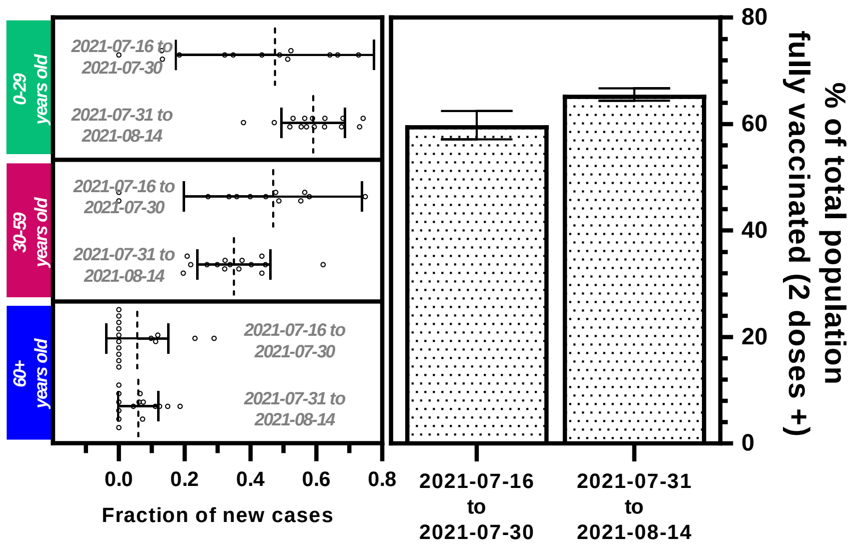


Figure S3: Comparison of the demographic distribution of new cases, per age group, and the proportion of the total population which was fully immunized (2 doses +) during the 15-day period preceding and following the onset of the Delta (B.1.617.2) variant of concern in the community.

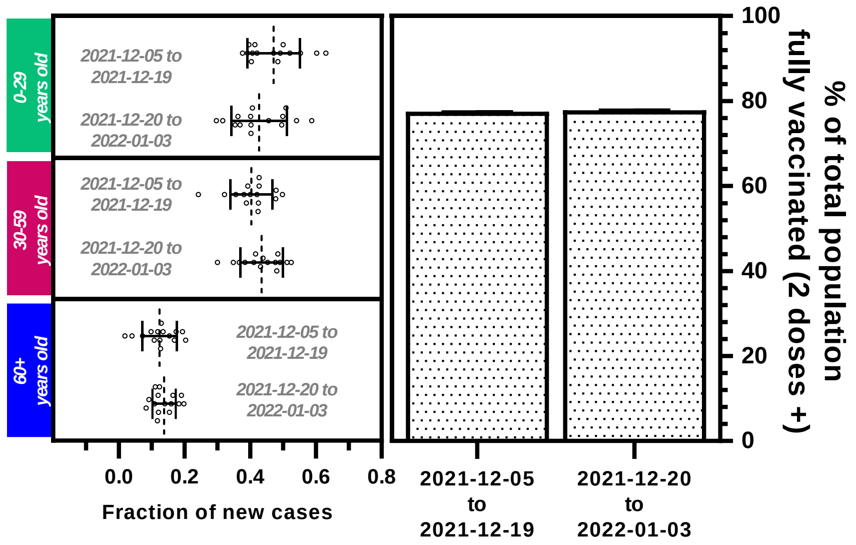


Figure S4: Comparison of the demographic distribution of new cases, per age group, and the proportion of the total population which was fully immunized (2 doses +) during the 15-day period preceding and following the onset of the Omicron (B.1.1.529) variant of concern in the community.

42

### Appendix A

Detailed methodologies for RT-qPCR analysis are included below:

**Ottawa**

Singleplex RT-qPCR quantification of the SARS-CoV-2 viral signal was performed by targeting the N1 and N2 gene regions [6,7]. Singleplex RT-qPCR quantification of the pepper mild mottle virus (PMMoV) concentration was also performed on the same samples. The TaqMan Fast Virus 1-Step Master Mix (Applied Biosystems, CAT# 4444436, USA) was utilized for all RT-qPCR reactions in this study. Every sample was investigated for inhibition via dilution experiments, and the samples were run in technical triplicates. Quantification was performed via 5-6 point standard curves using the EDX SARS-CoV-2 standard RNA material (Exact Diagnostics, CAT# COV019, USA). Samples that met the following quality control criteria were included for data analysis: i) standard curves are linear ($R^2 \geq 0.95$); ii) the copies/well occurred in the linear range of the standard curve, and iii) the primer efficiency was between 90%‑120%. Additionally, samples, where technical replicates had Ct values ≥ 0.5 standard deviations of the mean, were excluded from the analysis. 3 µL and 1.5 µL of RNA template/standard were loaded into each qPCR reaction (N1 & N2, and PMMoV, respectively), and primers and probes were present at concentrations of 500 nM and 125 nM, respectively, for a final reaction volume of 10 µL. The copies of SARS-CoV-2 (N1/N2) detected per reaction were normalized against quantified copies of PMMoV from corresponding samples. The assay limit of detection (ALOD; ≥95% detect.) was previously assessed and determined to be approximately 2 copies/reaction for both N1 and N2, and approximately 3 copies/reaction for PMMoV [7,8]. The assay limit of quantification (ALOQ; CV = 35%) was determined to be approximately 3.2 copies/reaction for N1, approximately 8.1 copies/reaction for N2, and approximately 7.6 copies/reaction for PMMoV. Evaluation of viral recovery efficiency was also performed according to published protocols (D'Aoust et al. 2020a); with the data in this study not being corrected for recovery efficiency. Non-template controls and positive controls were included in every RT-qPCR run, and extraction blanks were also performed daily. All probes and primers utilized in this study are presented in the Supplemental Material (Table S2).

**Toronto**

Singleplex RT-qPCR quantification of the SARS-CoV-2 viral signal was performed by targeting the N1 and N2 gene regions. Singleplex RT-qPCR quantification of PMMoV concentration was also performed on the same samples. Quantification was performed via 5 to 6 point standard curves using a custom, a synthetic linearized plasmid containing the complete N1/N2 genes and partial PMMoV gene. Standards were diluted in poly-A carrier solution. A 4-fold serial dilution of standards with concentrations from 3.9 to 1.6 x $10^4$ copies/reaction was applied for N1 and N2. Since PMMoV is present at a much higher concentration in RNA from wastewater, serial dilution from 16 to 2 x $10^6$ copies/reaction was included in the standard curve for this target. The TaqMan Fast Virus 1-Step Master Mix (Applied Biosystems, CAT# 4444436, USA) was utilized for all RT-qPCR reactions in this study. Samples that met the following quality control criteria were included for data analysis: 1) standard curves are linear ($R^2 \geq 0.99$); 2) the primer efficiency was between 95%-105%. 4 µL of RNA template/standard were loaded into each qPCR reaction, and primers and probes were present at concentrations of 500 nM and 125 nM, respectively, for a final reaction volume of 10 µL. Standard curves for each target, three negative controls (no template control, whole process control, and carrier control), and two positive controls (with concentrations similar to the tested samples) were run along with samples on each 384 PCR plate. All quantifications for each target were made using standard curves that were run on the same plate. N1 and N2 were run with triplicates and PMMoV was run with duplicates. The limit of quantification (LOQ) was estimated to be the lowest concentration on the standard curve, which was 3.94 copies/reaction for N1 and N2, and 16 copies/reaction for PMMoV, respectively. The limit of detection (LOD) was set at 1 copy/reaction (theoretical number) when all negative controls were negative. If there was any amplification in negative controls greater than 1 copy/reaction, the LOD was estimated to be the average of all the negative controls. Samples with no amplification or quantification lower than 0.5 copies/reaction (half of the theoretical LOD) were replaced with 0.5 copies/reaction and flagged as below LOD. A 10-fold dilution of each RNA sample was also included for PMMoV, and quantification was compared with the PMMoV in the original RNA sample to check for inhibition. If the ratio of PMMoV in the original sample to the 10x diluted sample was less than 5, all targets were run again after being diluted. All probes and primers utilized in this study are presented in the Supplemental Material (Table S1).

**Kitchener, Peel and Waterloo**

Quantification of SARS-CoV-2 N gene (N1, N2, N63-Universal), D63G (Delta specific), and PMMoV in the extracted RNA was performed using the TaqPath™ 1-Step RT-qPCR Master Mix, CG (Applied Biosystems, CAT# A15299, USA) on CFX Connect Real-Time PCR or CFX Opus 96 Real-Time PCR systems (Bio-Rad, Hercules, USA). Quantification was performed via 5–6-point standard curves using the EDX SARS-CoV-2 standard RNA material (Exact Diagnostics, CAT# COV019, USA). PMMoV and HCoV-229E were quantified using dsDNA gblocks (Integrated DNA Technologies, Ottawa, Canada). Sequences are listed in Supplemental Materials (Table S1). All qPCR plates were run with no template controls, no reverse transcriptase controls, extraction blanks, and a positive control. RT-qPCR reactions were run in triplicate using 5 µL RNA template and a final reaction volume of 20 µL. The PCR conditions were as follow: RT at 50°C for 15 min, 95°C for 2 min, 45 cycles of 95°C for 3 sec followed by 60°C (for N2, 229E, Zebrafish, and MS2) or 55°C (N1, PPMoV, D63, and D63G) for 30 sec. In addition to these targets, samples were tested for inhibition in the PCR reaction using a multiplexed zebrafish and MS2 assay. RNA extracts were plated using a mastermix spiked with zebrafish dsDNA gblock and MS2 RNA fragment. Amplification of the zebrafish and MS2 sequences in the samples were compared to the amplification of sequence in qPCR water. If amplification in the sample was +/- 1 Ct of the amplification in qPCR water, the samples were considered to be inhibited. All probes and primers utilized in this study are presented in the Supplemental Material (Table S1).

**Edmonton and Calgary**

SARS-CoV-2 detection and quantification were performed by one-step RT-qPCR as previously described [9]. The TaqMan Fast Virus 1-Step Master Mix (Applied Biosystems, CAT# 4444436, USA) was utilized for all RT-qPCR reactions in this study. 5 µL of RNA template was used in a total reaction volume of 10 µL. Samples were tested in duplicate for each SARS-CoV-2 qPCR target and were considered positive if at least 2 out of the total 4 RT-qPCR reactions tested positive. PMMoV was also detected and quantified in each sample to normalize the SARS-CoV-2 concentration in wastewater. All RT-qPCR runs included positive and negative controls for quality control purposes. Virus quantification was performed using a standard curve prepared from a series of 10-fold dilutions (1.66 to $1.66 \times 10^6$ copies per reaction) of an RNA standard. The RNA standard was prepared via in vitro transcription from long oligonucleotide sequences (gblocks) carrying the E-gene target of SARS-CoV-2 (Integrated DNA Technologies, Ottawa, Canada) and titrated with Nanodrop™ [10]. As the efficiency of RT-qPCR for SARS-CoV-2 E, N1, N2 and PMMoV were similar, the same standard curve for the E gene was used for quantification of all three targets. The limit of detection (LOD) of the one-step RT-qPCR assay for SARS-CoV-2 was 1.6 copies/PCR or 80 copies/100 mL of wastewater water and the efficiency of the standard curve was 99%. The coefficient of variation for Ct values from 20 replicates of RT-qPCR was 1.48% for $1.66 \times 10^3$ copies/reaction and 1.76% for $1.66 \times 10^5$ copies/reaction, respectively [9].

#### RT-qPCR quantification of the B.1.1.7 (Alpha) VOC in wastewater

In all locations where the B.1.1.7 (Alpha) VOC allelic was surveyed (Ottawa, Toronto, Kitchener, Peel, Waterloo, Edmonton and Calgary), the detection and quantification of the allelic proportions of the B.1.1.7 (Alpha) VOC (based on the ND3L mutation) were performed as described by Graber *et al. (2021)*. All probes and primers utilized in this study are presented in the Supplemental Material (Table S1).

#### RT-qPCR quantification of the B.1.617.2 (Delta) VOC in wastewater

In all tested locations where the B.1.617.2 (Delta) VOC was surveyed (Ottawa, Kitchener, Peel and Waterloo), the detection and quantification of the allelic proportions of the B.1.617.2 (Delta) VOC (based on the D63G mutation) was performed as described in the section 2.2 (Kitchener, Peel, Waterloo) All probes and primers utilized in this study are presented in the Supplemental Material (Table S1).

#### RT-qPCR quantification of the B.1.1.529 (Omicron) VOC in wastewater

In the tested location where the B.1.1.529 (Omicron) VOC was surveyed (Ottawa), the detection and quantification of the allelic proportions of the B.1.1.529 (Omicron) VOC (based on the P13L mutation) were performed as described in Appendix A.

**Description of P13L (Omicron-specific) RT-qPCR assay and use**

Allele-specific (P13L / C28311T) and universal (N1) RT-qPCR assays were performed in parallel to detect BA.1/BA.2/BA.3/C.37 variants and total SARS-CoV-2 signal respectively. Specific primers and N1 probe were used at a final concentration of 500 and 125 nM respectively. Sequences of the probes/primers can be found above in Supplemental Table 2. RT-qPCR reactions were run in triplicate and cycling was performed on a CFX Connect qPCR thermocycler (Bio-Rad, Hercules, CA) as follows: 50°C for 5 minutes, 95°C for 20 seconds, and 45 cycles of 95°C for 3 seconds, then 55°C for 45s. No-template controls (NTC) showed either no amplification after 45 cycles or, rarely, poor amplification above 40 cycles. Reactions were considered positive when Ct<40. The N1-universal assay is non-specific to any lineage but measures the quantity of SARS-CoV-2 in the same region as the P13L assay. Proportion (%) of Omicron is calculated as per the equation below:

$$Proportion\ (\%)\ of\ signal\ attributable\ to\ Omicron\ variant = \left(\frac{2^{-C28311T}}{2^{-N1}}\right) * 100\%$$

This qPCR assay was developed to target the mutation P13L on the N gene, which is unique to the BA.1/BA.2/BA.3/C.37 variants. During the study period C28311T was found at a frequency of >99% in SARS-CoV-2 genomes assigned to omicron lineages and deposited to GISAID and <1% in other genomes (covidcg.org).

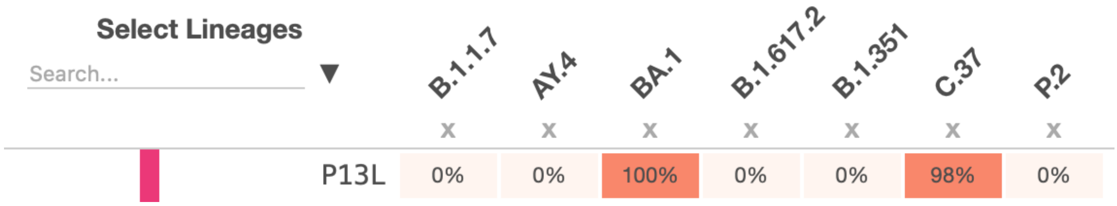

The assay is shown to be specific to Omicron/Lambda VOCs via standard curve testing, showing little to no cross-reactivity using Twist Bioscience’s #48 standard (Twist BioScience, San Francisco, USA).
